# Prism adaptation treatment for upper-limb Complex Regional Pain Syndrome: a double-blind randomized controlled trial

**DOI:** 10.1101/2020.03.18.20038109

**Authors:** Monika Halicka, Axel D. Vittersø, Hayley McCullough, Andreas Goebel, Leila Heelas, Michael J. Proulx, Janet H. Bultitude

**Affiliations:** Centre for Pain Research, University of Bath, Bath, United Kingdom; Department of Psychology, University of Bath, Bath, United Kingdom; Department of Sport and Health Sciences, University of Exeter, Exeter, United Kingdom; Pain Research Institute, Department of Translational Medicine, University of Liverpool, Liverpool, United Kingdom; Department of Pain Medicine, Walton Centre NHS Foundation Trust, Liverpool, United Kingdom; Optimise Pain Rehabilitation Unit, Oxford University Hospitals NHS Trust, Oxford, United Kingdom; Centre for Real & Virtual Environments Augmentation Labs, Department of Computer Science, University of Bath, Bath, United Kingdom

## Abstract

Initial evidence suggested that people with Complex Regional Pain Syndrome (CRPS) have reduced attention to the affected side of their body and the surrounding space, which might be related to pain and other clinical symptoms. Three previous unblinded, uncontrolled studies showed pain relief following treatment with prism adaptation, an intervention that has been used to counter lateralised attention bias in brain-lesioned patients. To provide a robust test of its effectiveness for CRPS, we conducted a double-blind randomized controlled trial of prism adaptation for unilateral upper-limb CRPS-I. Forty-nine eligible adults with CRPS were randomized to undergo two-weeks of twice-daily home-based prism adaptation treatment (n = 23) or sham treatment (n = 26). Outcomes were assessed in person four weeks prior to and immediately before treatment, and immediately after and four weeks post-treatment. Long-term postal follow-ups were conducted three and six months after treatment. We examined the effects of prism adaptation versus sham treatment on current pain intensity and CRPS symptom severity score (primary outcomes); as well as sensory, motor, and autonomic functions, self-reported psychological functioning, and experimentally tested neuropsychological functions (secondary outcomes). We found no evidence that primary or secondary outcomes differed between the prism adaptation and sham treatment groups when tested at either time point following treatment. Overall, CRPS severity significantly decreased over time for both groups, but we found no benefits of prism adaptation beyond sham treatment. Our findings do not support the efficacy of prism adaptation treatment for relieving upper-limb CRPS-I. This trial was prospectively registered (ISRCTN46828292).

## 1. Introduction

Complex Regional Pain Syndrome (CRPS) is associated with continuous pain in one or more limbs accompanied by sensory, motor, and autonomic disturbances that are disproportionate to any inciting injury [35]. Individuals with CRPS can also show neuropsychological symptoms reminiscent of hemispatial neglect after brain injury [33]. These can present as distorted cognitive representations of the CRPS-affected limb(s) [45,50,72,85,94], reduced attention to the affected limb(s) and corresponding side of external space [11,21,25,27,76,84], poorer mental representation of the affected side of space [99], and spatially-defined motor deficits [84]. The extent of these neuropsychological changes has been associated with the severity of clinical signs of CRPS [25,46,50,75,76,84,85,105] and could pertain to its central mechanisms [86].

Prism adaptation (PA) is a sensorimotor training technique used to reduce lateralised biases in attention, spatial representations, and (ocular)motor performance in hemispatial neglect after brain injury [55,69,90]. Considering similar neuropsychological deficits in CRPS, three previous studies tested the efficacy of PA in a total of 13 patients with this condition. They reported significant relief of pain and other CRPS symptoms following eight to 20 PA sessions performed with the affected arm when participants adapted towards their affected side [9,12,100]. The reduction in pain lasted up to two weeks. Thus, PA has the potential to durably relieve pain and other symptoms of CRPS. Because PA is quick (5-10 minutes a day), inexpensive, and self-administered, it is an appealing intervention compared to more intensive neurocognitive treatments like graded motor imagery [73]. However, the strength of available evidence for PA is limited, because it was only tested in small samples, without any control treatments or blinding.

The mechanisms through which PA could relieve pain are unclear. One possibility is that it increases attention to the CRPS-affected side relative to the unaffected side. Indeed, when one patient underwent adaptation in the opposite direction such that the theoretical attention bias away from the affected side would be exacerbated, their pain increased [100]. More severe self-reported “neglect-like” symptoms and spatial attention and motor biases have been related to greater pain intensity and worse long-term pain outcomes [25,84,85,105]. A potential second mechanism is that PA restores normal sensorimotor integration, the disruption of which is thought to contribute to pathological pain, including CRPS [8,38,63,100]. This is consistent with findings that individuals without spatial biases can also benefit from PA [12].

We conducted a double-blind, randomized, sham-controlled trial of PA for upper-limb CRPS-I. We hypothesised that two weeks of twice-daily PA treatment would reduce the primary outcomes of pain intensity and CRPS symptom severity more than sham treatment of the same intensity. We also predicted greater reductions in the secondary outcomes of neuropsychological symptoms (i.e. biases in spatial cognition, motor control, and body representation), clinical signs of CRPS, and self-reported CRPS-related and psychological disturbances following PA compared to sham treatment. The outcomes were assessed at six time points: to establish a one-month pre-treatment baseline, and to examine any immediate effects of PA and their retention at one, three, and six months post-treatment.

## 2. Methods

### 2.1. Study design and participants

The study was a two-arm parallel group RCT. It was prospectively registered (ISRCTN46828292 [42]) and the full details of the study are reported in the study protocol and analysis plan [34]. Any protocol deviations are specified in the relevant sections of the article. Anonymised participant-level data generated during the current trial (https://osf.io/ba6fq/), digital study materials (training protocol and video, PsychoPy experiment files and stimuli; https://osf.io/7fk2v/), and analysis scripts (https://osf.io/w67rx/) are publicly available in an Open Science Framework repository. The study was approved by the UK National Health Service (NHS) Oxfordshire Research Ethics Committee A and Health Research Authority (reference 12/SC/0557).

We aimed to address the following research questions:

1. Is two weeks of twice-daily PA treatment more effective in reducing pain and CRPS symptom severity than sham treatment?
2. Are there any improvements in other clinical signs of CRPS, psychological functioning, and neuropsychological symptoms following PA treatment?
3. How long are any benefits sustained for after the cessation of PA treatment?
4. Are there factors that can predict the CRPS progression over time and/or the response to PA treatment?
5. Are the neuropsychological abnormalities in CRPS (as compared to pain-free controls) related to clinical signs of CRPS?

Participants with CRPS were primarily recruited for the current RCT of PA treatment (questions 1-4), but we also collected measures of spatial cognition, motor control, and body representation at baseline (RS1) for comparison with pain-free controls and correlational analysis with clinical signs of CRPS (question 5; data reported elsewhere [32]).

Recruitment was conducted via post through the National CRPS-UK Registry, internal registries of the Royal United Hospitals and Walton Centre NHS Foundation Trusts, and clinicians’ referrals through the Oxford University Hospitals NHS Foundation Trust and other NHS pain clinics in the UK. Word of mouth, print and online advertisements, as well as social media were further used to disseminate information about the study. Participants were recruited between March 2017 and December 2018, and the final long-term follow-up took place in July 2019.

Following provisional assessment of eligibility through a phone interview, recruited participants took part in four research sessions (RS) at the University of Bath (n = 33), University of Liverpool (n = 9), or in the participant’s home (for participants who were unable to travel; n = 7). Participants gave written informed consent at the beginning of RS1, prior to any study-related procedures. The research sessions involved in-person assessment of eligibility criteria and of the primary and secondary outcomes, including self-report questionnaires, clinical assessments, and tests of neuropsychological functions. Each RS lasted from two to four hours, including breaks between the assessments. The data collection schedule is presented in Figure 1. The baseline was measured over two research sessions (RS1 and RS2) separated by four weeks. Immediately after RS2, participants commenced a two-week home-based treatment period. Treatment outcomes were measured over two research sessions, one immediately (RS3) and one four weeks (RS4) after completing the treatment. Two long-term follow-ups were conducted via post – one at 12 weeks (LTFU1) and one at 24 weeks (LTFU2) after completing the treatment. The flow of participants through each stage of the study is displayed in a CONSORT diagram (Figure 2).

**Figure 1.**
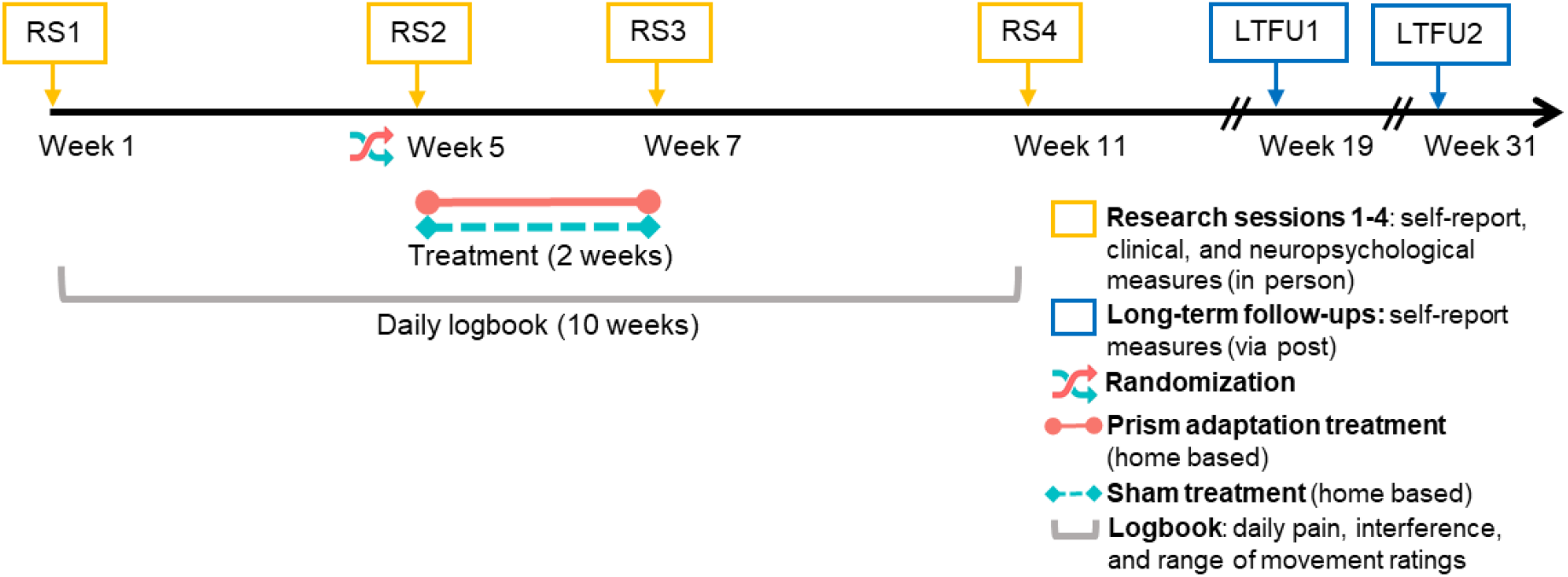
Schedule of data collection and interventions.

**Figure 2.**
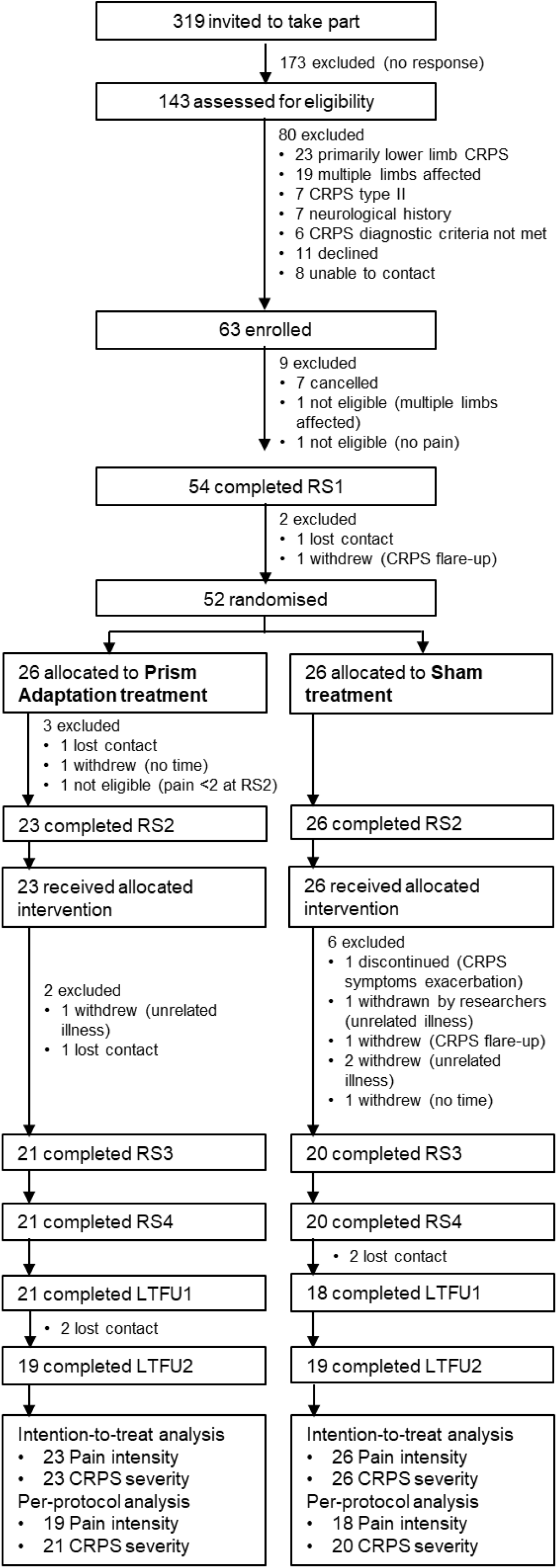
CONSORT diagram. Flow of participants through the study. RS1, research session 1; RS2, research session 2; RS3, research session 3; RS4, research session 4; LTFU1, long-term follow-up 1; LTFU2, long-term follow-up 2; Intention-to-treat analysis, participants who received allocated intervention; Per-protocol analysis, participants who completed allocated intervention, RS3-4 (CRPS severity), and LTFU1-2 (Pain intensity). Note that three participants who were allocated to Prism adaptation treatment did not attend RS2 or did not meet the eligibility criteria in RS2, thus they were not trained and did not receive any treatment, and were not included in the intention-to-treat analysis.

Participant inclusion criteria were: being aged 18-80 years; having a diagnosis of CRPS-I primarily affecting one upper limb based on the Budapest research criteria [35]; having a CRPS diagnosis for ≥3 months at the time of RS1; and having a current pain intensity ≥2 on a 0-10 Numeric Rating Scale. Exclusion criteria were: lacking sufficient English language ability to provide informed consent; being classified as legally blind; reporting a history of neurological disorder (e.g. stroke, neurodegenerative disease, or traumatic brain injury); having CRPS in the opposite limb meeting the Budapest clinical or research criteria; reporting confirmed nerve damage (CRPS-II); reporting or showing dystonia or other physical impairment that would prevent satisfactory execution of PA/sham treatment; or reporting severe psychiatric comorbidity (e.g. schizophrenia [102]) that could be associated with perceptual changes. Inclusion and exclusion criteria were assessed in RS1 and RS2.

### 2.2. Interventions

Both groups were instructed to continue any usual treatments (including medications) but were asked not to change their treatment regimens throughout the duration of the trial if possible. Current treatments and any changes are reported in Supplemental Table S1.

#### 2.2.1. Prism adaptation treatment

Participants randomised to the PA treatment used welding goggles fitted with 35-diopter Fresnel lenses that induced approximately 19° lateral optical deviation (visual shift) away from the CRPS-affected side. In each treatment session, participants were seated approximately 50 cm from a wall or other vertical surface (the actual distance was adjusted individually to correspond to the participant’s almost fully extended arm, thus it differs from the 60 cm distance anticipated in the trial protocol [34]). An A4 sheet was positioned on the wall in a landscape orientation at eye-level and in line with their body midline. There were two targets (2 cm-diameter red circles) on the pointing sheet, located 12.5 cm to the left and 12.5 cm to the right of participant’s body midline. While wearing the prism goggles, participants used their CRPS-affected arm to perform 50 pointing movements, as fast as possible, alternating between the left and right target.

An example of prism adaptation is illustrated in Figure 3. Prismatic shifts were directed away from the CRPS-affected side, thus participants with left-CRPS would use rightward-shifting prism goggles as illustrated in the figure. Due to the rightward visual shift, pointing would initially err to the right. However, with repeated movement execution and motor learning, the pointing would become increasingly accurate, as the movements would adjust in the opposite direction (to the left). This adaptive realignment of sensorimotor reference frames [82,101] would produce movement after-effects towards the left (affected) side. That is, once the goggles were removed, if participants were to point to the target again, their pointing would temporarily err to the left. Conversely, participants with right-CRPS would use leftward-shifting prismatic goggles to induce adaptive realignment (movement after-effects) towards their affected side. Studies from neurologically healthy individuals and stroke patients show that these short-term movement after-effects are accompanied by a longer-lasting realignment of attention, spatial representations, and lateralised (ocular)motor performance in the same direction as the after-effect [3,14,24,43,47,53– 55,65,66,69,89,90,95,96,98,104].

**Figure 3.**
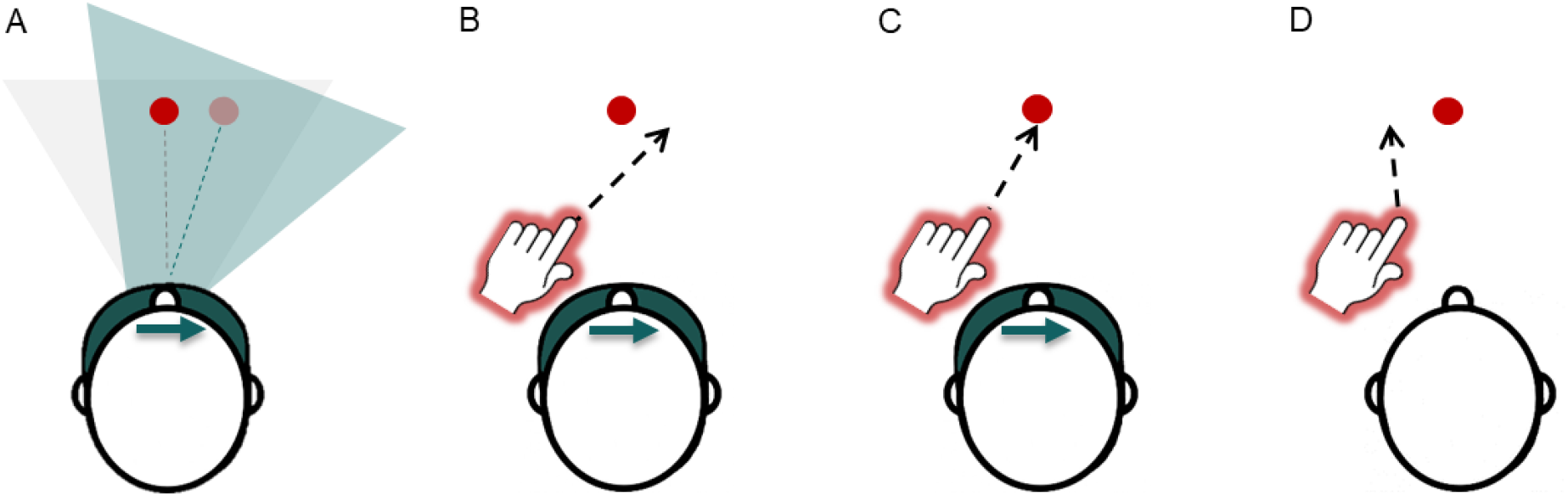
Prism adaptation procedure. In this example, participant with left-CRPS is using rightward-shifting prisms (A-C), which induce adaptation towards the left (affected) side. For clarity of illustration, only one target (red circle) is represented in the figure. However, the treatment procedure involved two targets presented in the left and right side of space, and participants’ pointing movements alternated between the left and right targets. (A) Prism goggles shift visual image to the right. Blue triangle represents a shift of visual perspective and perceived target location (pale red circle), relative to real location of the target (light grey triangle, dark red circle). (B) Pointing movements initially err to the right. (C) Adaptive realignment results in correct pointing movements. (D) Goggles are removed and pointing movements err to the left (after-effect).

The chosen direction of PA, inducing a visual shift away from the affected side and thereby an after-effect towards the affected side, is consistent with previous PA studies in CRPS [9,12,100] and the technique’s application in rehabilitation of hemispatial neglect after brain injury [55,90]. To enhance the effects of PA, welding goggles occluded the first half of the arm movement and participants were encouraged to point as quickly as possible. Both of these measures are thought to reduce any deliberate mis-aiming on behalf of the participants and encourage greater adaptive realignment (i.e. “true” sensorimotor adaptation) [19,82,83]. Previous studies in hemispatial neglect and CRPS demonstrated that the chosen number of movements (50 per session) is sufficient to induce adaptation measured as pointing after-effects, and changes in spatial cognition [9,90,100]. Note that the immediate movement after-effects were not measured in this trial.

Immediately after RS2, participants were trained in person in how to carry out the treatment by a research psychologist JHB or ADV (neither of whom were involved in any data collection) according to a standardised protocol (available in study materials). Once the researcher was satisfied that the participant understood the treatment procedure, they performed the first treatment during this training session under the guidance of the researcher. At the end of the training session participants received a pair of prism goggles in a sealed opaque bag, a pointing sheet, written instructions, and a link to a video tutorial to take home. In addition to the treatment that they underwent during training, participants were instructed to perform twice-daily self-guided treatment sessions at home for two weeks, resulting in 29 treatment sessions in total. The number of sessions per day and days of treatment were based on regimens that have previously been shown to reduce hemispatial neglect following stroke [23,24,47,69,95,96]. This regimen was also more intense than those used in previous studies demonstrating CRPS reduction following PA treatment [9,12,100], however we also considered that it would not be too much of a burden for participants. They were instructed to commence the home-based treatment on the day following RS2, perform one session in the morning and on in the evening, and record the start and end time of each session in a provided logbook.

#### 2.2.2. Sham treatment

Participants randomized to the sham treatment carried out exactly the same procedure as described above, except they used welding goggles fitted with neutral lenses that did not induce any lateral visual shift [10,69]. The neutral lenses distorted the acuity and clarity of vision to a similar extent as prism lenses (only without any lateral shift), therefore the two treatment arms were similar aside from the sensorimotor adaptation.

### 2.3. Randomisation and blinding

Participant randomization was performed 1-5 days before RS2 by JHB, who was not involved in any data collection. Participants were randomly assigned to either PA or sham treatment group with equal allocation ratio, using MINIM [68] software to minimize baseline (RS1) group differences in current pain intensity, CRPS severity score, primarily affected arm, pre-CRPS dominant hand, sex, age, presence of CRPS in other body parts, presence of other non-CRPS pain, and CRPS duration. The primary outcome measures (current pain intensity and CRPS severity score) were given double weighting compared to the other minimisation characteristics as we considered matching the two groups for these factors to be a higher priority. Note that this allocation procedure was updated in the trial registration [42] after the first two participants were enrolled and provided RS1 data, but before they were allocated to treatment. The update reflects the use of MINIM software instead of blocked minimization to automate the allocation procedure. As per the trial protocol [34], participants excluded between treatment allocation and RS3 (Figure 2) were removed from the minimization procedure so that subsequently recruited participants could be allocated independent of these exclusions and according to the current pool of participants remaining in each arm.

The only researchers who were aware of individual treatment allocations were those who randomised the participants and/or trained them in carrying out PA or sham treatments and provided them with prism or neutral goggles (JHB and/or ADV). These researchers were not involved in the assessments of any outcomes at any point in the trial. In RS3, the participants returned the goggles in a sealed opaque bag to MH, which she handed unopened to JHB. The researcher responsible for enrolment and all data collection (MH) remained blinded to participants’ treatment allocation until the last participant completed their RS4. Following RS4, there were no further in-person assessments as the long-term follow-up was conducted via postal questionnaires and scored by blinded research assistants. The participants were blinded to their treatment allocations throughout the entire duration of the trial. They were informed that they might receive real or sham treatment, and that both involved reaching out to touch visual targets with their affected arm while wearing goggles that distort vision. However, participants were not made aware of the specific nature of the intervention nor the differences between the types of goggles used in the two treatment arms. All documentation and instructions referred to the treatment arm as “sensorimotor training”.

### 2.4. Measures

#### 2.4.1. Demographics

In RS1, participants reported on demographic characteristics, including age, sex, and handedness prior to CRPS onset. They completed two versions of Edinburgh Handedness Inventory [78]: rating their recalled (prior to CRPS onset) and current hand preference. Total scores can range from −100 (extreme left-handedness) to 100 (extreme right-handedness). To approximate the functional impact of CRPS, we calculated an absolute difference between current and recalled handedness scores, that is, change in handedness. We also interviewed the participants regarding their clinical history, including the date and type of any inciting injury, CRPS duration (time since diagnosis), any co-morbidities, and any ongoing treatments for CRPS.

#### 2.4.2. Primary outcomes

The primary research question was (1) whether two weeks of twice-daily PA treatment is more effective in reducing pain and CRPS symptom severity than sham treatment. Change between RS2 and RS3 in current pain intensity and CRPS symptom severity score were the primary outcomes. In RS1-RS4 and LTFU1-LTFU2, participants rated their current pain intensity in the CRPS-affected limb on a Numerical Rating Scale from 0 (no pain) to 10 (pain as bad as you can imagine). This measure was taken from the Brief Pain Inventory (item 6) [13], and has been recommended as a core outcome for chronic pain trials [17,31]. CRPS severity was assessed in RS1-RS4 according to a standardised protocol [34,37]. Eight self-reported symptoms and eight signs evaluated upon clinical examination were scored as 0 (absent) or 1 (present) based on sensory testing, and visual and manual examination. The summed CRPS severity score can range from 0 (no CRPS symptoms) to 16 (most severe CRPS symptoms). The CRPS severity score has good discrimination abilities, concurrent validity, and adequate sensitivity to change [36,37], and was recommended as the core outcome measure for CRPS clinical studies [31].

#### 2.4.3. Secondary outcomes

Our secondary research question was (2) whether there are any improvements in other clinical signs of CRPS, psychological functioning, and neuropsychological symptoms following PA treatment. Detailed description of and rationale for the secondary outcome measures is available in a published study protocol [34]. Below we provide the basic details of how these outcomes were quantified.

##### 2.4.3.1. Self-report measures

Self-reported secondary outcomes measured in RS1-RS4 and LTFU1-LTFU2 included questionnaires about pain, body representation, and emotional functioning. These were chosen based on recommendations for core outcome measures for chronic pain trials [17] and the existing literature on CRPS implicating other relevant measures (e.g. [30]). For pain-related outcomes, we used the Brief Pain Inventory (0-10 scale for each subscale; higher scores indicate greater pain intensity/interference [13]) and Pain Detect Questionnaire (-1-38 scale; higher scores indicate greater neuropathic component of experienced pain [26]). Body representation was measured using the Bath CRPS Body Perception Disturbance Scale (0-57 scale; higher scores indicate greater distortions [49]). For emotional functioning, we used Tampa Scale for Kinesiophobia (17-68 scale; higher scores indicate more severe pain-related fear of movement and re-injury [67]) and Profile of Mood States (17-229 scale; higher scores indicate greater mood disturbance [64]). In RS1, participants also completed Revised Life Orientation Test (0-24 scale; higher scores indicate higher optimism level [93]), and Patient Centred Outcomes Questionnaire (each item rated on 0-10 scale; higher scores indicate higher usual, desired, expected, and considered successful in terms of the treatment outcome levels of pain, fatigue, emotional distress, and interference, and higher importance of improvement in each of these areas [87]). These two measures were included to assess whether two treatment groups were matched on their average optimism and expectations of outcomes, because these factors can affect the success of novel treatments [4,51,103]. In post-treatment RS3-4 and LTFU1-LTFU2, participants rated their impression of how much their activity limitations, symptoms, emotions, and overall quality of life related to CRPS changed due to treatment, using the Patient Global Impression of Change questionnaire (1-7 scale; 1 indicates no change or worsening of symptoms; higher scores indicate greater improvement [40]).

Throughout the first 10 weeks of the trial (RS1-RS4), participants rated their average level (over the past 24 hours) of pain intensity, the degree to which their symptoms interfered with their daily life, and range of movement in the affected limb, using daily logbooks (0-10 Numerical Rating Scales; higher scores indicate greater pain intensity, symptoms interference, and better range of movement), to track the precise time course of any changes on these outcomes.

##### 2.4.3.2. Clinical assessments

In RS1-RS4 we assessed participants’ CRPS signs and symptoms to determine whether the Budapest research criteria were met and to calculate the CRPS severity score. In addition to in-person assessments, photographs and videos of both limbs were double-scored for the presence of colour asymmetry, dystrophic changes, and motor abnormalities by a trained research assistant who was blind to treatment allocation, affected limb, and time point of assessment. Cohen’s kappa statistics for inter-rater agreement were significantly different from zero, indicating fair agreement for colour asymmetry (*κ* = .21, *p* = .004) and dystrophic changes (*κ* = .23, *p* < .001), and borderline slight/fair agreement for motor impairment (*κ* = .20, *p* < .001). We also objectively quantified sensory, autonomic, and motor functions. Sensory tests were performed on the most painful site on the CRPS-affected limb and the corresponding site on the unaffected limb (assessed first), unless specified otherwise.

Secondary outcomes of sensory function of the affected relative to unaffected limb included elements of quantitative sensory testing administered according to the standardised protocol [34,88]. Specifically, we measured Mechanical Detection Thresholds using von Frey filaments. A positive threshold ratio [(affected-unaffected)/affected] indicates increased tactile detection threshold (hypoesthesia) on the affected side. We measured Mechanical Pain Thresholds using pinprick stimulators. A positive threshold ratio [(unaffected-affected)/unaffected] indicates decreased pain threshold (hyperalgesia) on the affected side. Allodynia was assessed using a cotton ball, a Q-tip, and a brush. An arithmetic mean of 15 ratings for these sensations from 0 (no sharp, pricking, stinging, or burning sensation) to 100 (most intense pain sensation imaginable) quantifies the severity of allodynia on the affected limb. We also measured Two-Point Discrimination thresholds on participants’ index fingertips according to a staircase procedure using a disk with one and two plastic tips separated by 2-15mm distance. A positive threshold ratio [(affected-unaffected)/affected] indicates higher tactile discrimination threshold (less precise discrimination ability) on the affected hand.

In addition to contributing to the CRPS severity score, the following measures were used as secondary outcomes of autonomic and motor function of the affected relative to unaffected limb: temperature difference (affected–unaffected; a negative score indicates that the affected limb was colder; absolute values were also analysed); oedema (affected–unaffected; higher scores indicate greater swelling of the affected limb); grip strength (affected/unaffected; scores <1 indicate weaker strength of the affected hand); and delta finger-to-palm distance (affected/unaffected; scores <1 indicate lower range of movement of the affected hand).

##### 2.4.3.3. Tests of neuropsychological functions

In RS1-RS4, the participants completed six experimental tests of the following neuropsychological functions: visuospatial attention in near space (Temporal Order Judgement, Landmark, and Greyscales tasks); mental representation of space (Mental Number Line Bisection task); spatially-defined motor function; and body representation (Hand Laterality Recognition task). A comprehensive battery of sensitive tests of distinct aspects of spatial cognition, motor control, and body representation deemed appropriate to fully capture any neuropsychological biases, how they would be affected by PA, and how they would relate to any changes in pain and other CRPS symptoms. Below we summarise how the neuropsychological functions were measured and quantified, whereas detailed descriptions of the experimental materials and procedures can be found in the trial protocol [34].

All experimental tasks were programmed and administered using PsychoPy software [79]. Those involving presentation of visual stimuli on a computer screen used a 34.5cm x 19.4cm touchscreen positioned at 50cm viewing distance. In all tasks (except the Mental Number Line Bisection), participants used a chinrest and fixated on a cross aligned with their body midline. When a manual response was required, participants used their unaffected hand to press the buttons, which were aligned orthogonally to the required response format (i.e. for left/right responses, participants pressed colour-coded bottom/top buttons). A short practice session was completed before each task. Data for stimuli/responses in the left and right sides of space for all tasks were recoded after collection in terms of affected and unaffected space relative to each participant’s CRPS-affected side.

The Temporal Order Judgement task measures covert spatial attention. Participants saw pairs of brief, identical light flashes, presented with different temporal offsets (±10-240ms range) onto a white table surface, one on each side of space. In one block participants reported which of the two lights they perceived first by saying “left” or “right”. In another block they reported which light they perceived second. The order of response type (first or second) was counterbalanced and results were averaged across these to account for any response bias [22]. We calculated the Point of Subjective Simultaneity, which expresses by how many milliseconds the light in the affected side of space had to precede (negative score) or follow (positive score) the light in the unaffected side of space for both lights to be perceived as simultaneous. Information that receives greater attention is perceived earlier than information that receives lesser attention [97]. Thus, a negative Point of Subjective Simultaneity indicates lower attention to the affected side of near space relative to the unaffected side.

The Landmark task [57] measures the visual representation of relative horizontal distance in near space. Participants saw pairs of landmarks (white circles) presented simultaneously, one on each side of space. While the distance between two landmarks was constant across all trials, their relative distance from the central fixation cross varied by 0.1° increments from ±8.1° to ±6.9° to the left and to the right. Participants indicated via a button press whether the left or the right landmark appeared closer to or further from the fixation cross. Results were averaged across two separate response blocks to account for any response bias. We calculated the Point of Subjective Equality, which expresses the relative distance (°) at which the landmark on the affected side of space had to be further from (negative score) or closer to (positive score) the fixation cross in order to perceive the two landmarks to be equidistant. A negative Point of Subjective Equality value indicates underestimation of the distance on the affected relative to the unaffected side, and thus underrepresentation of the affected side of near space.

The Greyscales task [77] measures overt spatial attention. Participants saw pairs of horizontal bars filled with greyscales presented one above the other. The two bars were mirror images of each other so that one bar was darker on the left side and the other bar was darker on the right side. Participants pressed a button to indicate which bar appeared to be darker overall. A negative value of the calculated index of spatial bias indicates that a higher proportion of overall darkness judgements was made based on the unaffected sides of the stimuli, consistent with lower attention to the affected relative to unaffected side.

The Mental Number Line Bisection task [99] measures mental representation of space, based on an implicit representation of numbers in a left-to-right linear arrangement [15]. The experimenter read aloud pairs of numbers that were separated by an interval of 9-64 digits. They were presented in ascending and descending order (e.g. 54 and 70, or 70 and 54) to account for any response bias. Participants were instructed to verbally report the subjective midpoint between the given pair of numbers, without making any calculations. A negative value of the calculated index of spatial bias is consistent with overestimating the subjective midpoint towards larger numbers (i.e. a rightward bias). Expressed relative to each participant’s CRPS-affected side, a negative index indicates a bias away from the affected side of the mental representation of space, or underrepresentation of the affected side of mental space.

The spatially-defined motor function task [59] measures directional hypokinesia and directional bradykinesia, that is slowing of initiation and execution of movements directed towards the affected relative to unaffected side. On each trial, participants held down a button with their finger until the target appeared either on the left or on the right side of a computer screen. Then participants were required to release the button, touch the target on the screen, and return their finger to the button as quickly as possible. Participants completed the task from three starting positions (button positioned to the left, right, or aligned with their body midline; order counterbalanced), alternating between their unaffected and affected hand, across six blocks in total. We used average movement initiation times (from the target onset to button release) and movement execution times (from the button release to touch on the computer screen) for each combination of hand starting position and target location to calculate indices of directional hypokinesia and bradykinesia towards the affected side [92], separately for each hand used to complete the task. Index A quantifies the speed of initiating/executing movements towards the affected side relative to the unaffected side. This index was calculated as: [central starting position (affected – unaffected target location) – affected starting position (affected – unaffected target location)]. Index A allows to dissociate motor and perceptual neglect (i.e. effect of target location), however, it involves movement trajectories of different length. Thus, we also derived Index B that directly quantifies the speed of initiating/executing movements of the same physical length towards the affected side relative to the unaffected side. Index B was calculated as: [central starting position (affected target location) – affected starting position (affected target location)]. Positive values of indices A and B indicate slowing of initiation/execution of movements directed towards the affected relative to unaffected side, suggestive of directional hypokinesia/bradykinesia towards the affected side.

The Hand Laterality Recognition task [94] measures body representation. Participants saw images of hands appearing either to the left or to the right of central fixation. The images depicted left or right hands in different postures and rotations from upright (0°, 90°, 180°, or 270°). Participants were required to indicate as fast and as accurately as possible (via a button press) whether each image depicted a left or a right hand. Accuracy rates and average reaction times to correctly responded-to trials for each depicted hand were averaged across two image locations, because the side of space effects were not the primary interest of this trial and will be reported elsewhere. We calculated two indices of hand laterality recognition as the differences between the depicted hands: accuracy index (unaffected hand – affected hand) and reaction time index (affected hand – unaffected hand). Positive scores indicate less accurate and slower recognition of depicted hands corresponding to participant’s affected hand, relative to depicted hands corresponding to their unaffected hand. Thus, positive accuracy and reaction time indices suggest distorted representation of the CRPS-affected limb.

### 2.5. Statistical analyses

#### 2.5.1. Sample size calculation

The study was powered to evaluate the effects of PA treatment on a change in the primary outcome of pain intensity between RS2 and RS3. We estimated [18] that a sample of 21 participants with CRPS per treatment group would provide 90% power to detect a minimal clinically significant reduction of 2 on the primary outcome of pain intensity (0-10 NRS; [20]), with a *SD* of 1.98 (based on our previous research [11]), and a 2-tailed alpha of 0.05. Although we aimed to obtain 42 complete data sets for RS1-RS4, one participant in the sham treatment group withdrew after we terminated recruitment, thus the total number of participants who completed these sessions equals 41.

#### 2.5.2. Analyses

The analysis plan can be found in the trial protocol [34], thus below we only describe the main steps, any details not previously specified, and any deviations from the protocol. We used IBM SPSS Statistics 25 [41], R 3.5.3 [81], and MATLAB 2018b [58] software to process and analyse the data. Data preparation procedures are reported in Supplemental Text S1. Throughout, we reported bootstrapped bias-corrected and accelerated 95% confidence intervals (BCa 95% CIs) around all mean and median values. We used bootstrapped *χ2* tests, bootstrapped *t*-tests (or their non-parametric alternatives in case of violation of parametric assumptions), and ANOVAs to compare mean values between treatment groups and between data collection time points. ANOVA is robust to moderate violations of normality and homogeneity of variance [6,7], and we used Greenhouse-Geisser corrections if the sphericity assumption was violated. However, where severe (i.e. more than borderline significant and in multiple conditions) violations of the assumptions of normality, homogeneity of variance, *and* sphericity were found, we used linear mixed models analyses with non-parametric bootstrapping procedures (n = 1000). For linear mixed models analyses, a model term made a significant contribution to predicting an outcome when the 95% CI around the coefficient estimate (*B*) did not include zero. For the remaining analyses, statistical significance was defined as *p* < .05. We used one-tailed tests for comparisons for which we had directional hypotheses (i.e. RS2 vs. RS3 comparisons, as we predicted greater reductions on the outcome measures in PA than sham treatment group), and two-tailed tests for the remaining comparisons. We controlled for type I errors in the primary (but not exploratory) analyses by using Holm-Bonferroni correction for multiple comparisons within analysis of each outcome and reported adjusted *p* values (*p*_*adj*_).

Our primary analysis involved the intention-to-treat population, that is, participants who received their allocated intervention (i.e. received in-person training immediately after RS2), regardless of their treatment adherence or completion of the outcome assessments (PA treatment = 23, sham treatment = 26). Note that the trial protocol defined this population as all participants allocated to treatment, which did not account for the possibility that they could withdraw before being trained in how to carry out their allocated intervention. This was the case for three participants (Figure 2) who were not included in the intention-to-treat sample as per an updated definition. In Supplemental Text S2 we report a supportive per-protocol analysis of those participants who provided complete outcome data and completed their allocated treatment (missed no more than six treatment sessions). The results of the per-protocol analysis are broadly consistent with the intention-to-treat analysis.

##### 2.5.2.1. Effects of PA treatment on the primary outcomes

To evaluate the effects of PA treatment on the first primary outcome of pain intensity (research question 1) and the time course of any changes (research question 3), we conducted a 2 (Group: PA treatment, sham treatment) x 6 (Time: RS1, RS2, RS3, RS4, LTFU1, LTFU2) ANOVA. We planned sixteen a-priori contrasts to compare RS1 vs RS2, RS2 vs RS3, RS3 vs RS4, RS2 vs RS4, RS2 vs LTFU1, RS4 vs LTFU1, LTFU1 vs LTFU2, and RS2 vs LTFU2 within each treatment group.

To evaluate the effects of PA treatment on the second primary outcome of the CRPS severity score (research question 1) and the time course of any changes (research question 3), we conducted a 2 (Group: PA treatment, sham treatment) x 4 (Time: RS1, RS2, RS3, RS4) ANOVA. We planned eight a-priori contrasts to compare RS1 vs RS2, RS2 vs RS3, RS3 vs RS4, and RS2 vs RS4 within each treatment group.

##### 2.5.2.2. Effects of PA treatment on the secondary outcomes

To evaluate the effects of PA treatment on self-reported pain and psychological functioning, sensory, motor, and autonomic function, and neuropsychological functions (research question 2), and the time course of any changes (research question 3), we conducted 2×6 and 2×4 ANOVAs and planned the same contrasts as described for the analyses of the primary outcomes.

##### 2.5.2.3. Predictors of the CRPS progression over time

To investigate whether any baseline factors could predict CRPS progression over time (research question 4), independent of the treatment, we used the data from the total sample (N = 49) to perform exploratory best subsets regression analyses on the overall change in pain intensity and CRPS severity score throughout the course of the study. This analysis differed from that proposed in the trial protocol in terms of (a) operationalisation of the pain outcome, (b) selection of potential predictors, and (c) regression model used. (a) Change on the primary outcomes was quantified as individual regression slopes fitted to each participant’s ratings of current pain intensity across RS1-LTFU2 (instead of planned RS1-RS4 comparisons, to capture change over a longer period) and to each participant’s CRPS severity scores across RS1-RS4. More negative slopes indicate greater improvement over time (i.e. reduction in pain and CRPS severity). (b) In the protocol, when specifying selection of potential predictors we planned to prioritize those factors on which participants with CRPS significantly differed from pain-free participants at baseline (research question 5; [34]). However, they showed no differences on the tests of neuropsychological functions (results reported elsewhere [32]). Thus, we instead included all primary and secondary outcomes as potential explanatory variables. That is, we included participants’ demographic characteristics, self-reported pain and psychological functioning, sensory, motor, and autonomic function, and neuropsychological functions, as measured in RS1. We restricted the pool of potential predictors by excluding factors that lacked linear relationships with each outcome or were collinear with other predictors (see Supplemental Text S1). (c) Instead of the planned linear mixed model regression, best subsets regression was deemed more appropriate to allow unbiased selection of the best combination of explanatory variables. Best subsets regression is an automated approach that performs an exhaustive search for the best subset of factors for predicting the outcome and returns the best model of each size (up to a specified number of predictors) [56]. Considering our sample size (N = 49), we compared best subsets models that included one up to five predictors of each outcome. From the five models, the one with the lowest Akaike Information Criterion was preferred as best fit. To address a potential issue of overfitting, we also performed a five-fold cross-validation [48] of each of the five models suggested by best subsets regression analyses. This approach randomly splits the data set into five folds (subsets of observations). Each model is trained using the 80% of the data (four folds) and then tested on the remaining 20% of the data (one fold). This process is repeated until each fold has served as a test subset. The average of errors recorded in each repetition is a cross-validation error. The lowest cross-validation error indicates best model performance.

## 3. Results

### 3.1. Participant characteristics

Table 1 presents baseline characteristics and comparisons between PA and sham treatment groups. On average, participants reported moderate pain intensity (6/10), comparable with previous studies on prism adaptation (5.8-6/10; [12,100]) and other neurocognitive treatments (5.3-7/10; [45,61,73]) for CRPS. Median CRPS severity score in our sample was higher than the average severity reported for individuals with stable CRPS in the validation study of this tool (13 vs. 11.2/16; [37]), possibly because we used stricter inclusion criteria (Budapest research diagnostic criteria; [35]). Our participants on average had longer CRPS duration compared to other studies of neurocognitive treatments for CRPS (58 vs. 5-24 months; [9,12,45,61,73,100]). The proportion of participants with CRPS affecting their right side of the body was consistent with a large population study [70], although it was lower than in small-sample studies on prism adaptation (41% vs. 71-80%; [12,100]). Both the mean age and proportion of females were consistent with those previously reported in CRPS [12,37,70,91,100]. The most common comorbidities in our participants were depression (37%), anxiety (22%), migraines (16%), fibromyalgia (14%), and asthma (14%). These conditions were found to be prevalent in CRPS in previous population studies [52,71]. The most common treatments in the current sample included weak or strong opioids (57%), anticonvulsants (47%), paracetamol (45%), antidepressants (45%), physio-, hydro-, or occupational therapy (39%), and nonsteroidal anti-inflammatory drugs (35%; see Supplemental Table S1). Overall, demographic and clinical characteristics of our sample appear to be representative of general population of people with CRPS [2,37,70,91] and comparable to those reported in previous research investigating neurocognitive treatments for CRPS [9,12,45,61,73,100], except for the longer average disease duration in our study.

**Table 1.** Baseline (RS1) participant characteristics by treatment group (intention-to-treat analysis)

| Measure | Prism adaptation<br>treatment (n = 23) | Sham treatment<br>(n = 26) | Contrast |
| --- | --- | --- | --- |
| **Minimisation factors** |  |  |  |
| Current pain intensity (/10) *M* | 5.96 [5.02, 6.80] | 6.15 [5.26, 7.00] | $t(47) = -0.33, p = .741, d = 0.10$ |
| CRPS severity score (/16) *Mdn* | 13.00 [12.07, 13.93] | 12.50 [11.00, 13.00] | $U = 287.50, p = .809, d = 0.07$ |
| Primarily affected arm (% right) | 48% | 35% | $\chi^2(1) = .88, p = .348, \phi = -0.13$ |
| Pre-CRPS dominant hand (%<br>right) | 91% | 92% | $\chi^2(1) = .16, p = .898, \phi = 0.02$ |
| Sex (% female) | 83% | 85% | $\chi^2(1) = .04, p = .850, \phi = -0.03$ |
| Age (years) *M* | 47.35 [43.20, 51.95] | 45.31 [39.85, 50.85] | $t(47) = 0.53, p = .601, d = -0.15$ |
| CRPS in other body parts (%<br>present) | 13% | 8% | $\chi^2(1) = .38, p = .537, \phi = -0.09$ |
| Other non-CRPS pain (% present) | 44% | 39% | $\chi^2(1) = .13, p = .721, \phi = -0.05$ |
| CRPS duration (months since<br>diagnosis) *M* | 61.26 [47.15, 75.12] | 52.31 [39.49, 66.35] | $t(47) = 0.84, p = .388, d = -0.24$ |
| **Other control measures** |  |  |  |
| Optimism (Revised Life<br>Orientation Test; /24) *M* | 13.00 [10.97, 15.07] | 12.31 [11.00, 13.61] | $t(47) = 0.59, p = .560, d = -0.17$ |
| Mood disturbance (Profile of<br>Mood States; /229) *M* | 94.81 [79.96, 109.93] | 84.22 [70.94, 98.08] | $t(47) = 0.97, p = .349, d = -0.28$ |
| Fear of movement (Tampa Scale<br>for Kinesiophobia; /68) *M* | 38.79 [35.45, 41.95] | 40.38 [37.17, 43.35] | $t(47) = -0.65, p = .502, d = 0.19$ |
| Number of logged treatment<br>sessions (/29) *Mdn* | 29.00 [28.54, 29.46] | 29.00 [28.55, 29.45] | $U = 297.00, p = .965, d = 0.01$ |

Bootstrapped bias-corrected and accelerated 95% confidence intervals are reported in square brackets, [BCa 95% CI].
There were no significant differences between groups on any measures.

The randomization procedure successfully equated the two treatment groups on the minimization factors (Table 1). The two groups were also matched on baseline mean levels of optimism, mood disturbance, fear of movement, and expectations and criteria for success of the treatment (there were no significant differences between PA and sham treatment groups on any of the Patient Centred Outcomes Questionnaire items, *U*s ≥ 212.00, *ps*_adj_ ≥ .27, *ds* ≤ 0.51).

Eight participants (16%) withdrew from the study following treatment allocation. They were excluded from per-protocol analysis (Supplemental Text S2), but their RS2 data was carried forward for the purpose of the primary intention-to-treat analysis. We compared their baseline (RS1) pain intensity and CRPS severity against confidence intervals around the mean pain intensity and CRPS severity score of participants who remained in the trial. Out of those who dropped out, five participants had greater pain intensity and four participants had greater CRPS severity compared to those who remained. However, the same or lower pain intensity and CRPS severity scores were found in another three and four participants who dropped out, respectively.

### 3.2. Treatment adherence and participant blinding

Twenty-one out of 23 participants (91%) in the PA treatment group and 20 out of 26 participants (77%) in the sham treatment group missed no more than six treatment sessions according to their logbooks (see Supplemental Table S1). Two participants in the PA and six participants in the sham treatment group missed more than six treatment sessions and/or did not provide post-treatment outcome data. No other deviations from the treatment protocol were identified among the remaining participants. The extent of exposure to treatment (i.e. average number of logged treatment sessions) was not significantly different between the two treatment groups (Table 1). The median recorded durations of the treatment sessions according to the participants’ logbook entries were 2min 25s in the PA group and 2min in the sham treatment group.

At the end of RS4, we asked each participant (N = 41) which treatment they thought they received. They could respond “real [PA]”, “sham”, or “no idea”. Similar proportions of participants in each group made correct (real: 12.2%; sham: 12.2%) or wrong (real: 12.2%; sham: 7.3%) guesses as to their actual treatment allocation, or responded that they had no idea (real: 26.8%; sham: 29.3%), *χ*^*2*^(2) = .52, *p* = .771, *Cramer’s V* = 0.11. Only 12% of participants in each group correctly guessed their treatment allocation, therefore participant blinding was successful.

### 3.3. Effects of PA treatment on the primary outcomes

Figure 4 illustrates any changes on the primary outcomes and their time course in each treatment group (research questions 1 and 3). Despite the PA group showing some reduction in the current pain intensity scores immediately after treatment (RS3; Figure 4a), the ANOVA did not reveal any significant main effects of Time, *F*(4.04, 189.81) = 1.82, *p* = 0.126, *η*^*2*^_*p*_= 0.04, or Group, *F*(1, 47) = 0.26, *p* = 0.615, *η*^*2*^_*p*_= 0.01, nor did it show any significant interaction between these factors *F*(4.04, 189.81) = 0.66, *p* = 0.624, *η*^*2*^_*p*_ = 0.01. This indicates that there were no significant changes in pain intensity over time in either treatment group. Thus, contrary to our hypothesis, PA treatment did not reduce pain intensity more than sham treatment.

**Figure 4.**
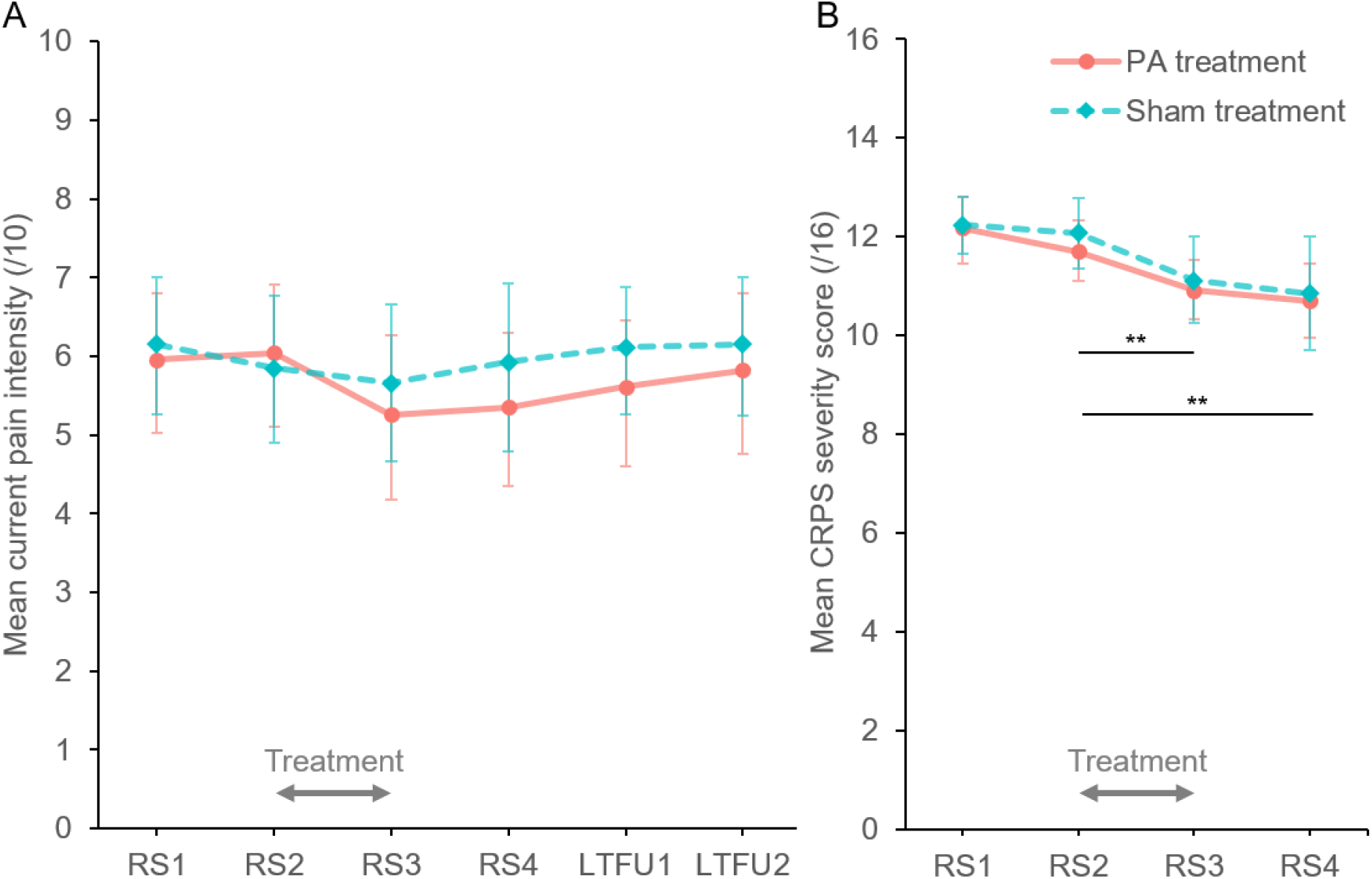
Primary outcomes (intention-to-treat analysis). Mean [BCa 95% CI] current pain intensity (A) and CRPS severity scores (B) in prism adaptation (PA; orange circles) and sham treatment (blue diamonds) groups in each time point. RS1, RS2, RS3, and RS4, research sessions 1, 2, 3, and 4; LTFU1 and LTFU2, long-term follow-up 1 and 2. Grey arrows indicate the treatment period. **Significant decrease in CRPS severity between RS2 and RS3, maintained at RS4, regardless of treatment, *ps*_*adj*_ < .01.

Analysis of the CRPS severity scores (Figure 4b) showed a large significant main effect of Time, *F*(2.28, 107.08) = 17.57, *p* < .001, *η*^*2*^_*p*_= 0.27, indicating that regardless of treatment, CRPS severity decreased over time (Figure 4b). Contrasts revealed a significant reduction in CRPS severity immediately after treatment (RS3; *Mdn* = 11.00, BCa 95% CI [11.00, 11.00]) compared to immediately before treatment (RS2; *Mdn* = 12.00, BCa 95% CI [12.00, 12.00]), *Z* = −3.91, *p*_*adj*_ = .002, *d* = 0.86. This reduction relative to RS2 was maintained four weeks after completing the treatment (RS4; *Mdn* = 11.00, BCa 95% CI [11.00, 11.00]), *Z* = −3.70, *p*_*adj*_ = .002, *d* = 0.81, but without further significant change from RS3, *Z* = −0.81, *p*_*adj*_ = .433, *d* = 0.16. CRPS severity did not change significantly between the first (RS1; *Mdn* = 13.00, BCa 95% CI [13.00, 13.00]) and the second baseline session, *Z* = −1.71, *p*_*adj*_ = .170, *d* = 0.35. There was no significant effect of Group, *F*(1, 47) = 0.17, *p* = .685, *η*^*2*^_*p*_< 0.01, nor was there any significant interaction effect, *F*(2.28, 107.08) = 0.17, *p* = .886, *η*^*2*^_*p*_ < 0.01, on the CRPS severity scores. Thus, contrary to our hypothesis, CRPS severity did not decrease more following PA compared to sham treatment, but both groups improved over the treatment period.

We compared mean changes in pain intensity and CRPS severity over the treatment period (RS3 – RS2) between PA and sham treatment groups. Effect sizes of these differences might be important for planning future studies. For current pain intensity, the effect size was small, *d* = 0.37, 95% CI [-0.20, 0.94]. Mean pain reduction in the PA treatment group was −0.78 points on 0-10 NRS scale, BCa 95% CI [-1.55, −0.15]. In the sham treatment group, mean pain reduction was −0.19 points, BCa 95% CI [-0.68, 0.28]. For CRPS severity score, the effect size was negligible, *d* = −0.13, 95% CI [-0.69, 0.43]. Mean CRPS severity reduction in the PA treatment group was −0.78 points on 0-16 scale, BCa 95% CI [-1.19, −0.38]. In the sham treatment group, the mean CRPS severity reduction was −0.96 points, BCa 95% CI [-1.54, −0.38]. On an individual level (Supplemental Figure S1), five participants in the PA group and four in the sham group achieved clinically significant reductions in pain (i.e. at least two-point decrease on 0-10 NRS scale [20]) over the treatment period. None of the participants achieved clinically significant reduction in CRPS severity (i.e. at least 4.9 points decrease on 0-16 scale, although this threshold is quite conservative [37]).

### 3.4. Effects of PA treatment on the secondary outcomes

Group average scores on the self-report questionnaires, clinical assessments, and neuropsychological tasks in each time point are reported in Table 2. Note that the confidence intervals around average baseline indices of the neuropsychological functions include zero, indicating that participants did not show significant biases in visuospatial attention or the mental representation of space, nor any differences in the recognition of the affected relative to unaffected hands. Furthermore, average indices of directional hypokinesia and bradykinesia vary between positive and negative values, suggesting that there were no systematic spatially-defined motor deficits at baseline. Complete results of a series of ANOVAs conducted to test the effects of PA on the secondary outcomes and their time course (research questions 2 and 3) are reported in the tables, and below we only refer to the effects directly relevant for our hypothesis, that is, Group x Time interactions.

**Table 2.**
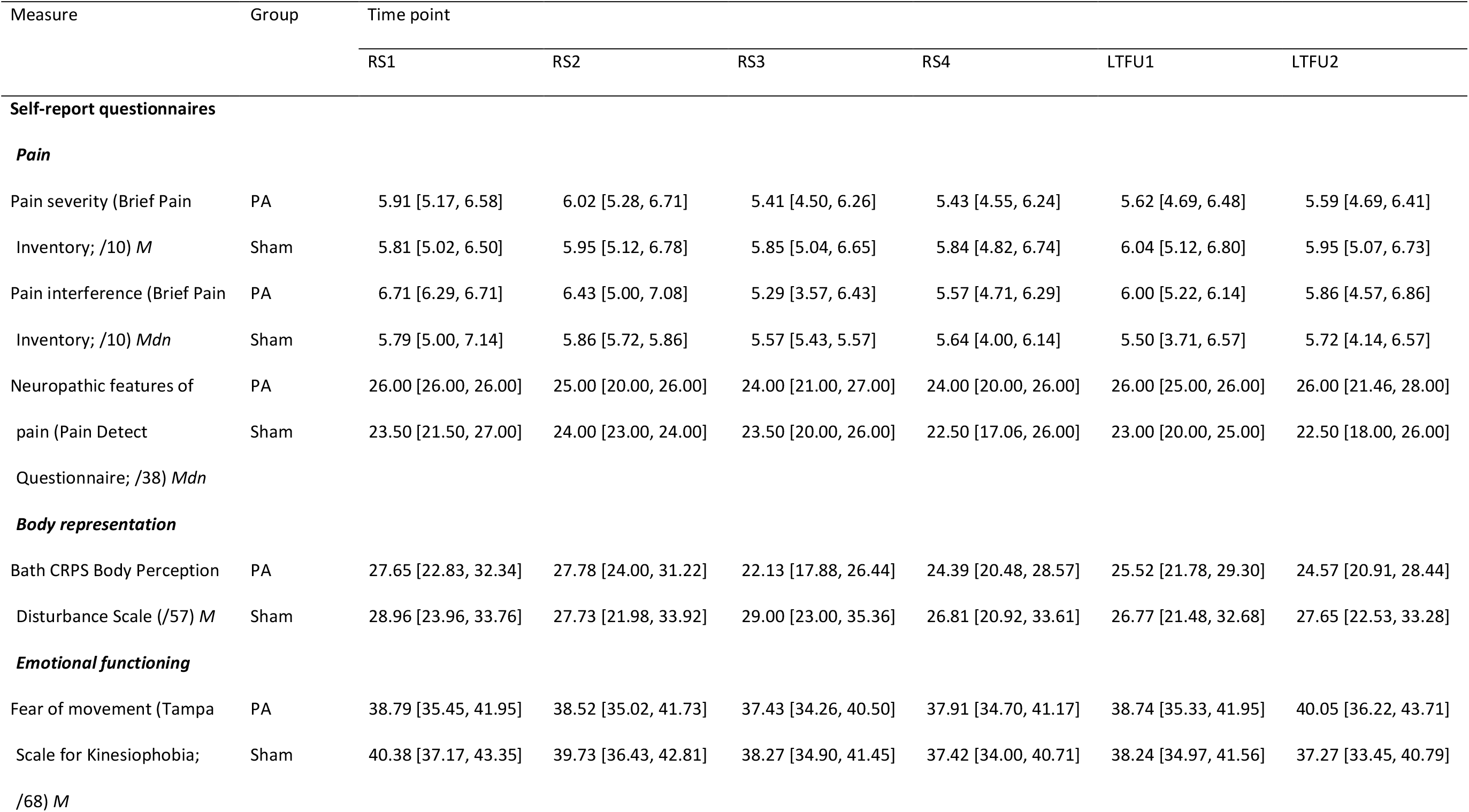
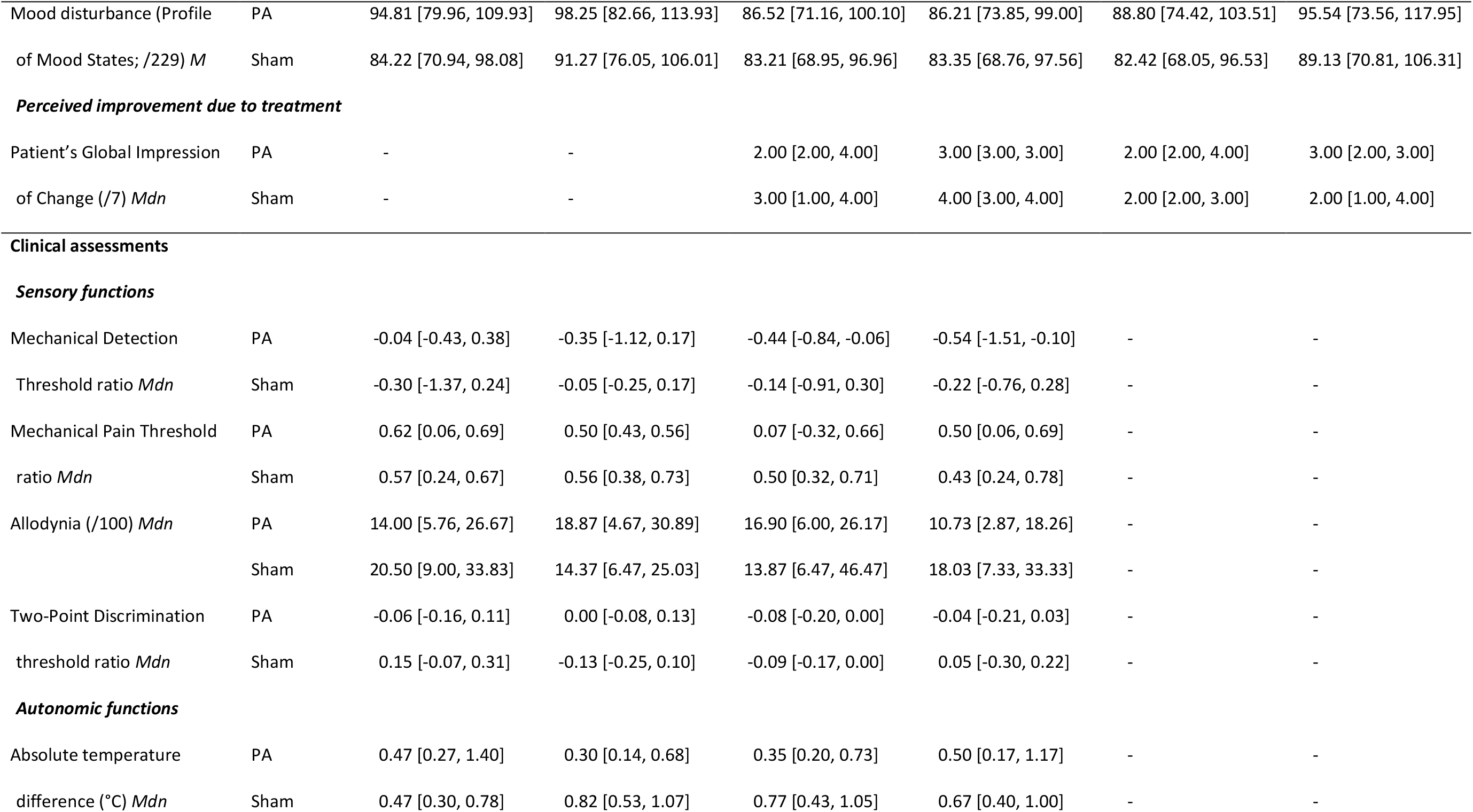
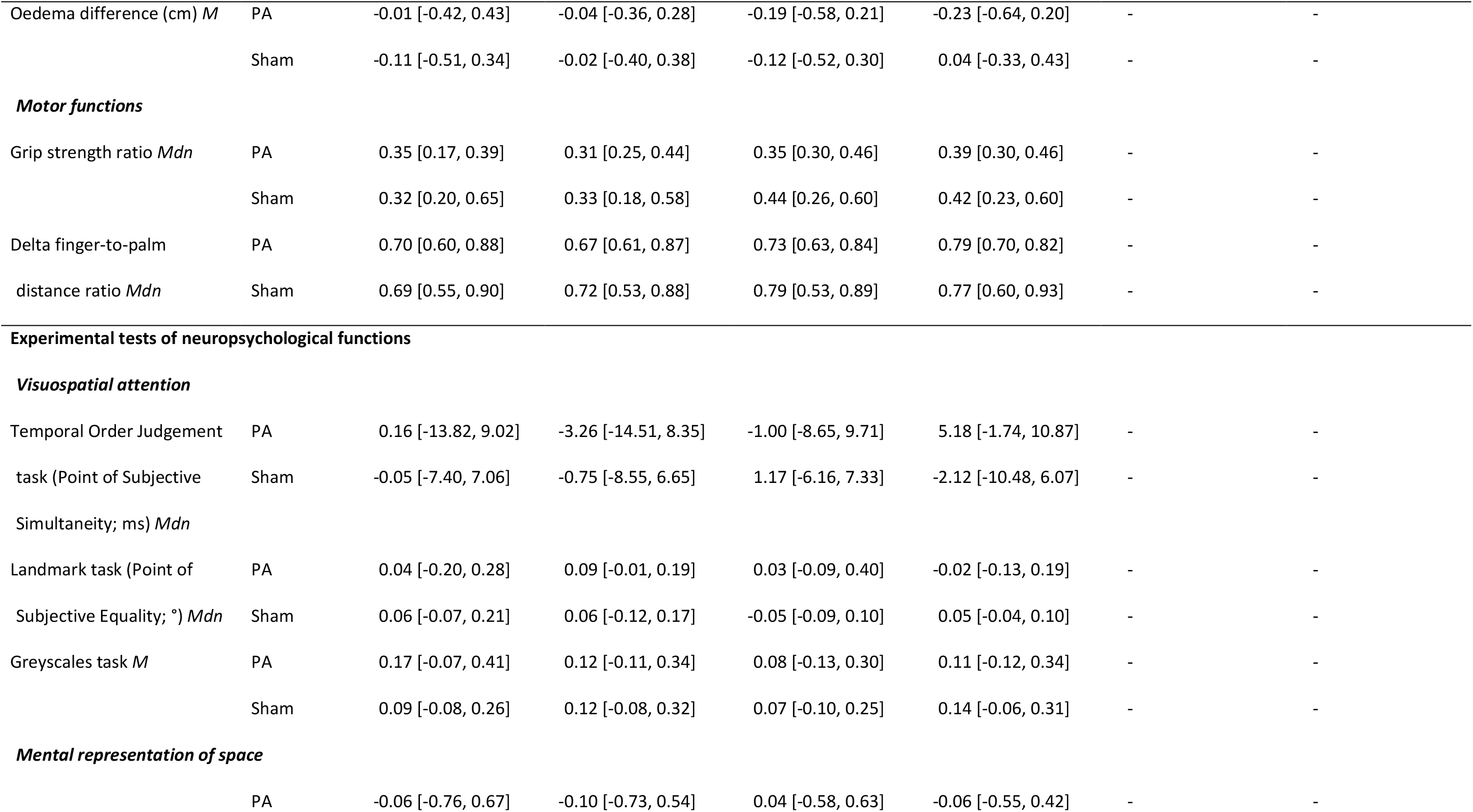
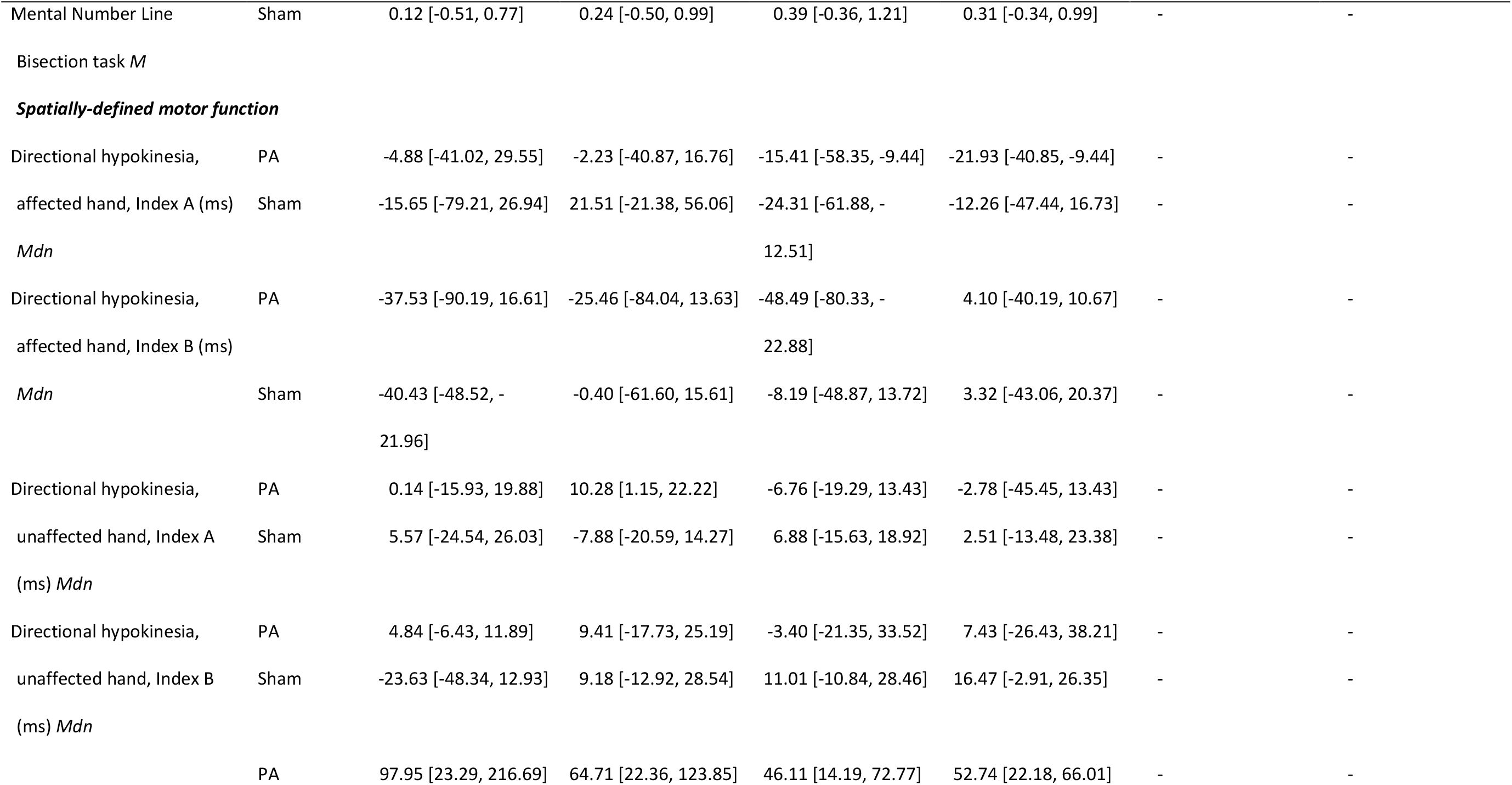
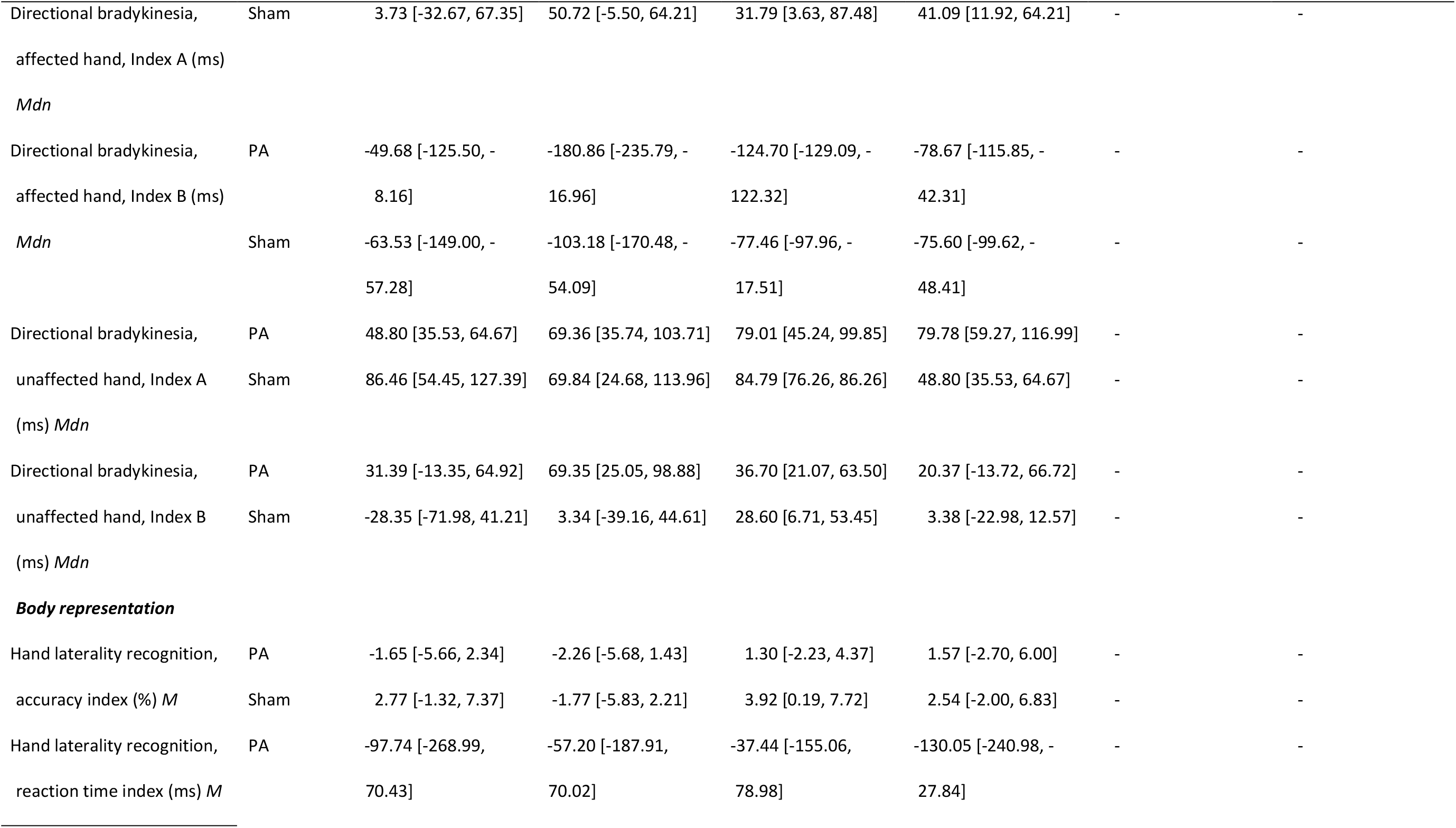
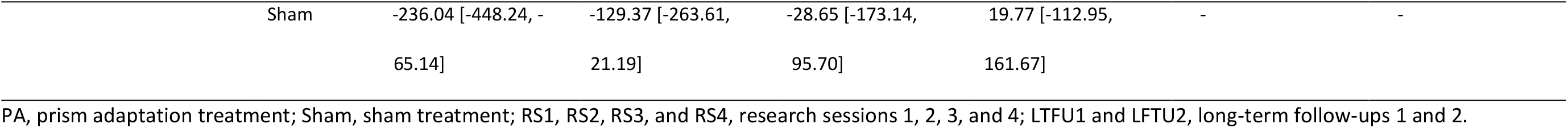
Mean or median values [BCa 95% CI] of self-reported; sensory, autonomic, and motor; and neuropsychological secondary outcome measures at each time point (intention-to-treat analysis)

Results of 2×6 ANOVAs on the self-reported pain-related, body representation, and emotional functioning outcomes, and 2×4 ANOVAs on the sensory, motor, autonomic, and neuropsychological functions, are reported in Table 3. Among these outcomes, the Mechanical Detection Threshold, Mechanical Pain Threshold, Two-Point Discrimination threshold, grip strength, and delta finger-to-palm distance ratios data, the Landmark task, and spatially-defined motor function data were analysed using linear mixed models regression due to severe violations of normality, homogeneity of variance, and/or sphericity assumptions. The results are reported in Table 4.

**Table 3.** Analysis of variance results for secondary outcome measures (intention-to-treat analysis)

| Measure | Effect | df<sup>†</sup> | F | p | $\eta^2_p$ |
| --- | --- | --- | --- | --- | --- |
| **Self-report questionnaires** |  |  |  |  |  |
| Pain severity (Brief Pain Inventory) | Time | 4.12, 193.81 | 1.24 | 0.295 | 0.03 |
|  | Group | 1, 47 | 0.19 | 0.664 | < 0.01 |
|  | Time x Group | 4.12, 193.81 | 1.06 | 0.379 | 0.02 |
| Pain interference (Brief Pain Inventory) | Time* | 2.88, 135.32 | 2.84 | 0.043 | 0.06 |
|  | Group | 1, 47 | 0.04 | 0.838 | < 0.01 |
|  | Time x Group | 2.88, 135.32 | 0.74 | 0.526 | 0.02 |
| Neuropathic features of pain (Pain Detect Questionnaire) | Time* | 3.29, 154.50 | 3.32 | 0.018 | 0.07 |
|  | Group | 1, 47 | 0.32 | 0.574 | 0.01 |
|  | Time x Group | 3.29, 154.50 | 0.61 | 0.625 | 0.01 |
| Bath CRPS Body Perception Disturbance Scale | Time | 3.41, 160.11 | 2.43 | 0.059 | 0.05 |
|  | Group | 1, 47 | 0.57 | 0.455 | 0.01 |
|  | Time x Group* | 3.41, 160.11 | 2.60 | 0.047 | 0.05 |
| Fear of movement (Tampa Scale for Kinesiophobia) | Time | 3.86, 181.61 | 2.41 | 0.053 | 0.05 |
|  | Group | 1, 47 | < 0.01 | 0.993 | < 0.01 |
|  | Time x Group* | 3.86, 181.61 | 2.89 | 0.025 | 0.06 |
| Mood disturbance (Profile of Mood States) | Time | 3.60, 169.21 | 2.29 | 0.069 | 0.05 |
|  | Group | 1, 47 | 0.36 | 0.554 | 0.01 |
|  | Time x Group | 3.60, 169.21 | 0.25 | 0.894 | 0.01 |
| Patient's Global Impression of Change | Time | 3, 120 | 0.96 | 0.414 | 0.02 |
|  | Group | 1, 40 | 0.02 | 0.890 | < 0.01 |
|  | Time x Group | 3, 120 | 0.56 | 0.644 | 0.01 |
| **Clinical assessments** |  |  |  |  |  |
| Allodynia (affected limb) | Time | 2.23, 104.67 | 1.03 | 0.367 | 0.02 |
|  | Group | 1, 47 | 0.25 | 0.616 | 0.01 |
|  | Time x Group | 2.23, 104.67 | 0.35 | 0.730 | 0.01 |
| Absolute temperature difference | Time | 3, 141 | 0.43 | 0.731 | 0.01 |
|  | Group | 1, 47 | 0.16 | 0.695 | < 0.01 |
|  | Time x Group | 3, 141 | 0.63 | 0.595 | 0.01 |

| Measure | Effect | *df*<sup>†</sup> | *F* | *p* | $\eta^2_p$ |
| --- | --- | --- | --- | --- | --- |
| Oedema difference | Time | 2.41, 113.08 | 0.99 | 0.387 | 0.02 |
|  | Group | 1, 47 | 0.06 | 0.805 | < 0.01 |
|  | Time x Group | 2.41, 113.08 | 1.86 | 0.153 | 0.04 |
| **Experimental tests of neuropsychological functions** |  |  |  |  |  |
| Temporal Order Judgement task (Point of Subjective Simultaneity) | Time | 1.70, 79.69 | 1.08 | 0.335 | 0.02 |
|  | Group | 1, 47 | 0.16 | 0.692 | < 0.01 |
|  | Time x Group | 1.70, 79.69 | 0.63 | 0.512 | 0.01 |
| Greyscales task | Time | 2.17, 101.82 | 0.57 | 0.581 | 0.01 |
|  | Group | 1, 47 | 0.02 | 0.899 | < 0.01 |
|  | Time x Group | 2.17, 101.82 | 0.52 | 0.609 | 0.01 |
| Mental Number Line Bisection task | Time | 2.39, 112.17 | 0.48 | 0.656 | 0.01 |
|  | Group | 1, 47 | 0.50 | 0.481 | 0.01 |
|  | Time x Group | 2.39, 112.17 | 0.14 | 0.899 | < 0.01 |
| Hand laterality recognition, Accuracy index | Time | 3, 141 | 2.39 | 0.072 | 0.05 |
|  | Group | 1, 47 | 1.54 | 0.221 | 0.03 |
|  | Time x Group | 3, 141 | 0.44 | 0.723 | 0.01 |
| Hand laterality recognition, Reaction time index | Time | 3, 141 | 1.32 | 0.269 | 0.03 |
|  | Group | 1, 47 | 0.05 | 0.826 | < 0.01 |
|  | Time x Group | 3, 141 | 1.48 | 0.224 | 0.03 |

\* Statistically significant effect ( $p < .05$ ).
† Greenhouse-Geisser adjusted degrees of freedom are reported where sphericity assumption was violated.

**Table 4.**
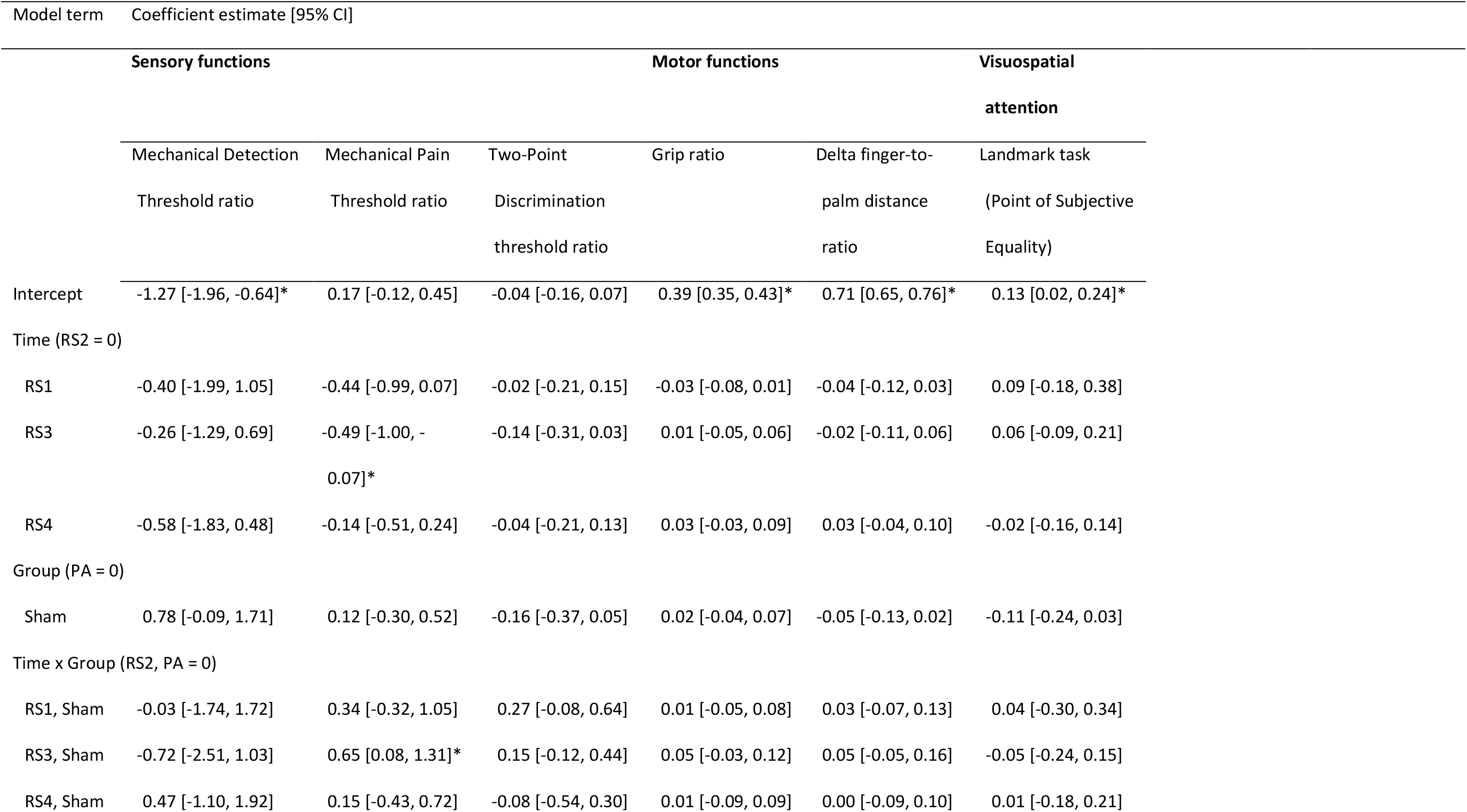
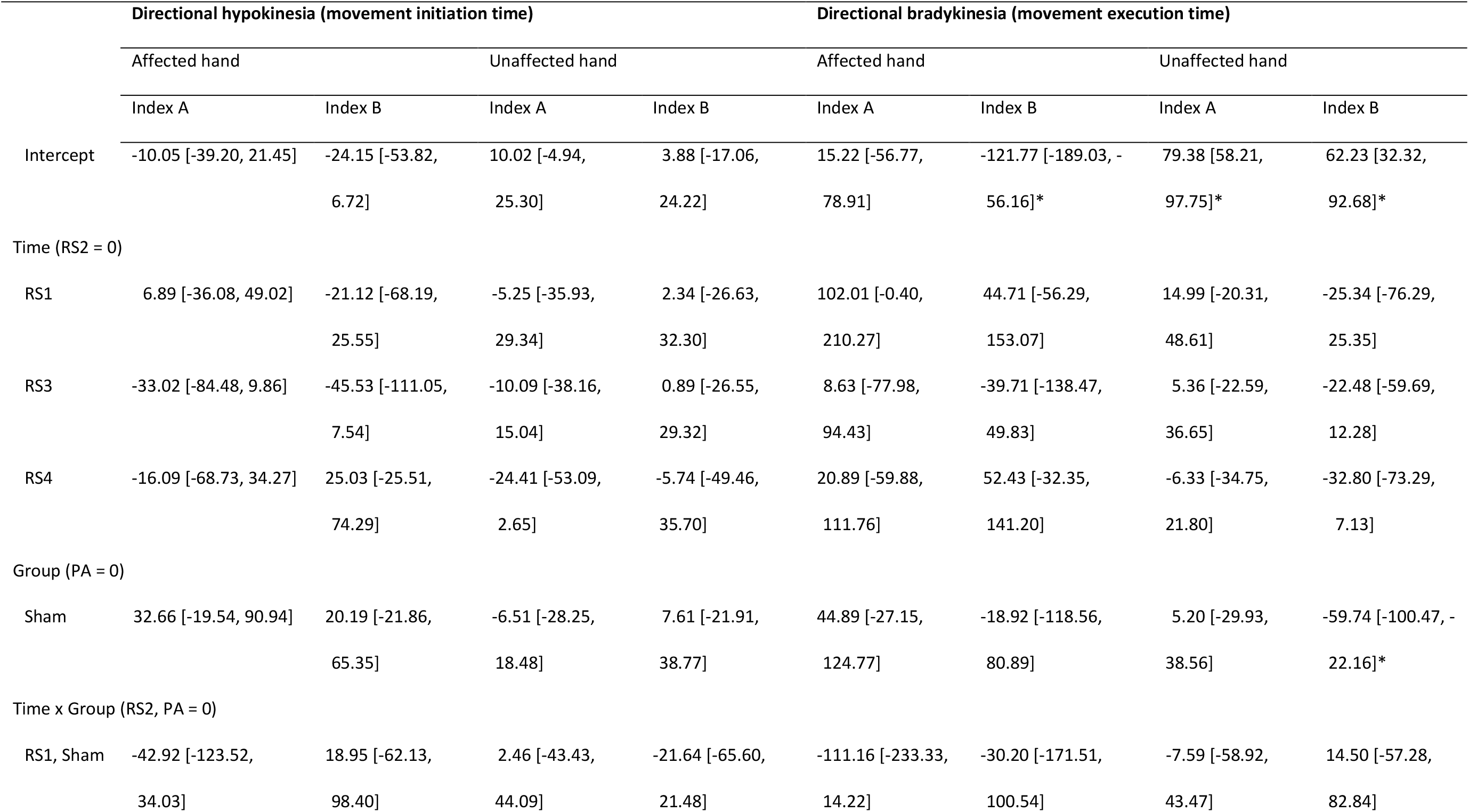
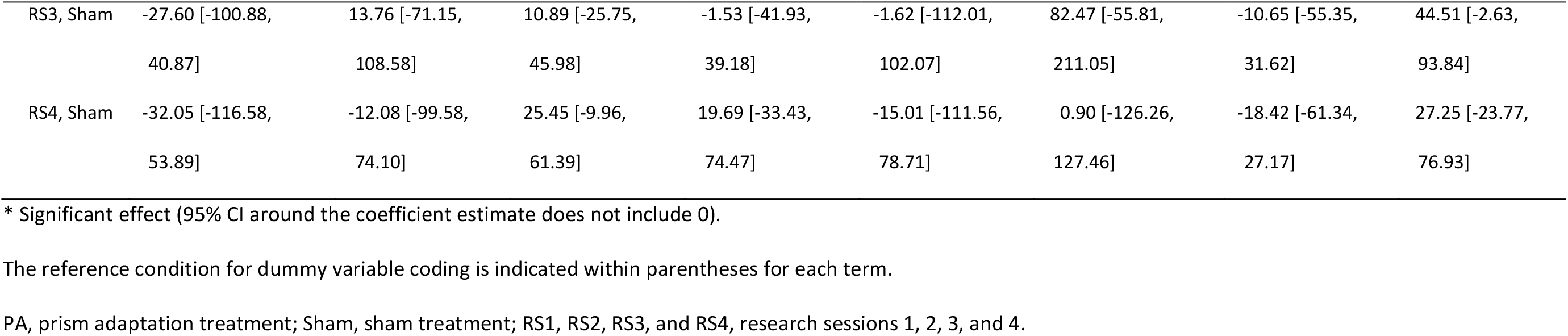
The results of the bootstrapped linear mixed models regressions of scores for the tests of sensory and motor function, visuospatial attention, and spatially-defined motor function (intention-to-treat analysis)

Contrary to our hypothesis, we found no evidence of significantly greater reductions in self-reported CRPS-related and psychological disturbances; or sensory, autonomic, and motor impairments following PA compared to sham treatment. We also did not find any significantly greater reductions in biases in spatial cognition, motor control, and body representation following PA compared to sham treatment. That is, most interactions between treatment group and time on these outcomes were not significant, and any effects revealed by the planned contrasts following the few significant interactions did not withstand correction for multiple comparisons. These effects are further elaborated below. A significant interaction on the Bath CRPS Body Perception Disturbance scores appeared to be due to reductions in body perception disturbance following the PA treatment, *ts* ≤ 2.86, *ps*_*adj*_ ≥ .336, *ds* ≤ 0.54. A significant interaction on the Tampa Scale for Kinesiophobia sores appeared to be driven by reductions in fear of movement following the sham treatment, *ts* ≤ 2.63, *ps*_*adj*_ ≥ .312, *ds* ≤ 0.26. A significant interaction on the Mechanical Pain Threshold ratios seemed to be due to a reduction in hyperalgesia over the treatment period in the PA group, *Zs* ≤ 1.68, *ps*_*adj*_ ≥ .440, *ds* ≤ 0.51. All these effects were no longer significant after Holm-Bonferroni correction and there were no other significant interactions. On average, participants in both treatment groups perceived their symptoms to be either “almost the same”, or “a little better” (2-3 out of 7 on the Patients’ Global Impression of Change) at each post-treatment time-point.

The PA and sham treatment groups also did not differ on their average daily logbook ratings of pain intensity, symptoms interference, and range of movement at any time point [pain intensity: *ts*(45) ≤ 1.75, *ps* ≥ .093, *ds* ≤ 0.51; symptom interference: *ts*(45) ≤ 1.24, *ps* ≥ .240, *ds* ≤ 0.36; range of movement: *ts*(45) ≤ 1.81, *ps* ≥ .062, *ds* ≤ 0.53]. The planned analyses of the number of days to reach peak improvement and from peak improvement to return to baseline on each of these measures [34] are not reported as they would not be informative in the absence of treatment effects, but logbook ratings for each group are illustrated in Supplemental Figure S2.

Overall, our analyses did not reveal any significant effects of PA compared to sham treatment on any of the secondary outcome measures.

### 3.5. Predictors of CRPS progression over time

Since there was no effect of treatment on the primary outcomes, we did not explore potential predictors of the response to PA treatment as proposed in the trial protocol [34]. However, to explore whether the absence of the PA effect could be explained by the lack of group-level neuropsychological deficits in our sample, we visualised individual relationships between the changes on the primary outcomes over the treatment period and baseline spatial bias and body representation distortion in Supplemental Figures S4 and S5, respectively. Overall, there were no apparent clusters of participants or relationships between these factors. Subgroup analyses of whether response to treatment depended on clinical phenotypes of CRPS [16] or baseline neuropsychological differences are also reported in Supplemental Text 3, showing results consistent with the primary analyses.

To address research question 4 about the predictors of CRPS progression over time, we explored which baseline factors (RS1) could predict overall change in pain intensity (across RS1-RS4 and LTFU1-LTFU2) and CRPS severity (across RS1-RS4). Table 5 summarises the models identified via best subsets regression analyses, and their respective values of our model selection criteria (the lowest Akaike Information Criteria and cross-validation errors indicating the best models). Greater reduction in pain intensity was best predicted by smaller change in hand preference since CRPS onset (absolute change on the Edinburgh Handedness Inventory; *t* = 2.34, *p* = .024, *ß* = 0.33) in a one-factor model, *F*(1, 46) = 5.46, *p* = .024. Greater reduction in CRPS severity was best predicted by lower pain intensity (*t* = 3.69, *p* < .001, *ß* = 0.52), less swelling of the affected limb (*t* = 2.52, *p* = .015, *ß* = 0.37), and more accurate recognition of images of the affected hand (i.e. smaller Hand laterality recognition accuracy index; *t* = 2.43, *p* = .019, *ß* = 0.32), as measured at baseline (RS1), in a three-factor model, *F*(3, 45) = 6.23, *p* = .001.

**Table 5.**
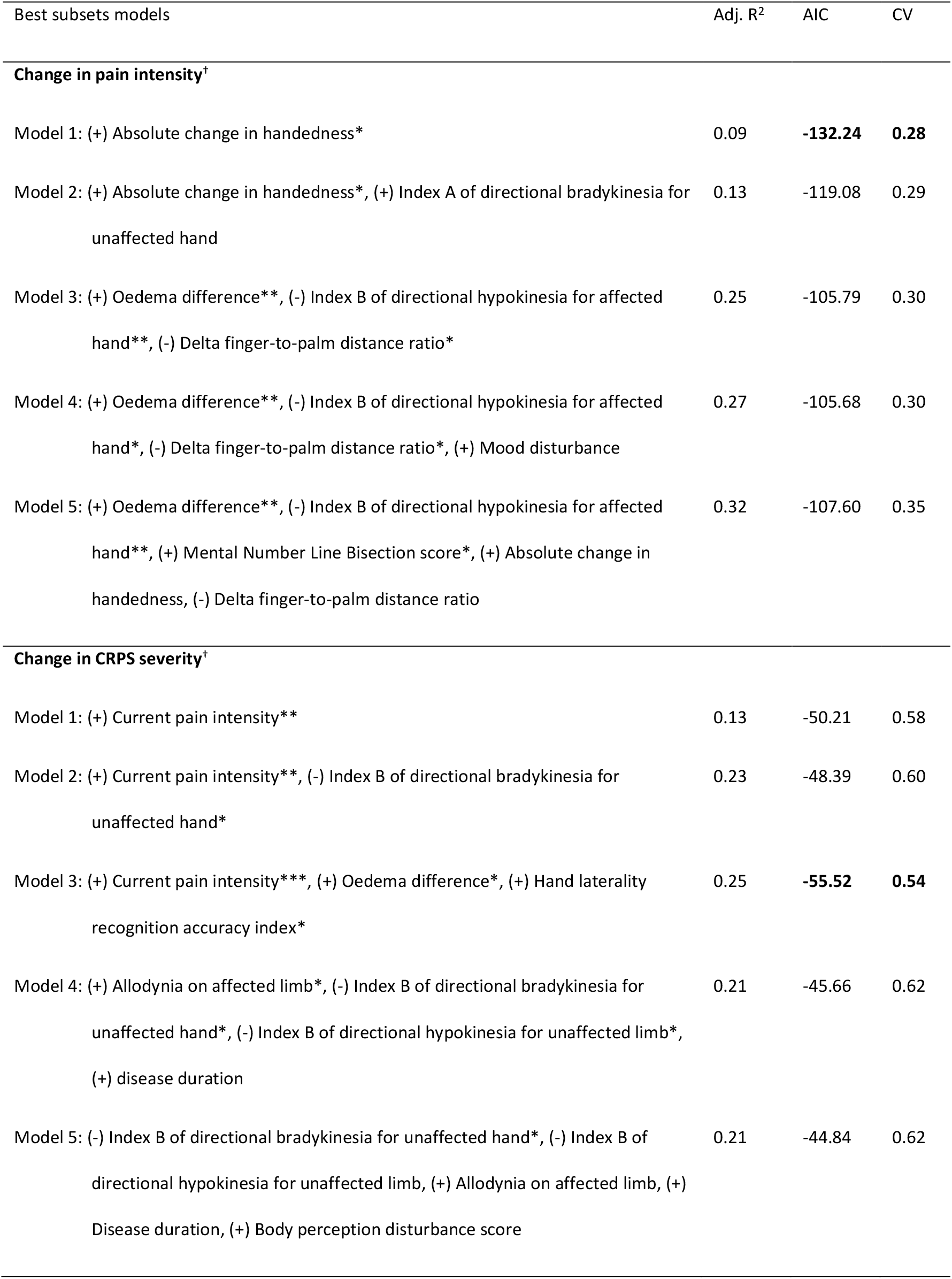
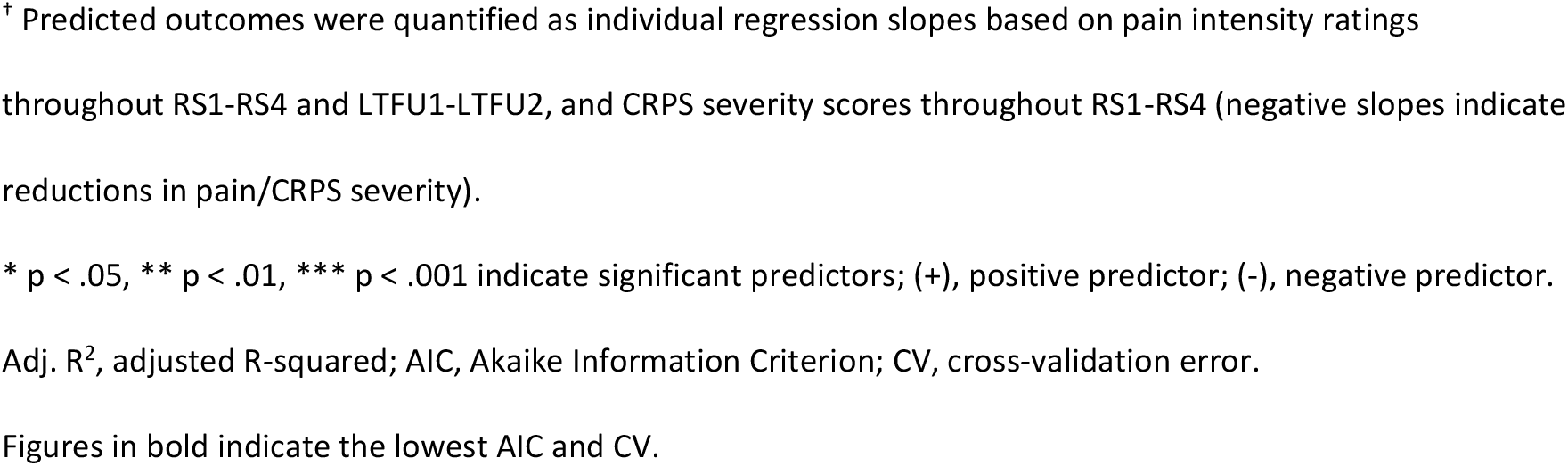
Best subsets of factors (as measured in RS1) for predicting overall change in pain intensity and CRPS severity throughout the study period (intention-to-treat analysis)

| Best subsets models | Adj. R<sup>2</sup> | AIC | CV |
| --- | --- | --- | --- |
| **Change in pain intensity<sup>†</sup>** |  |  |  |
| Model 1: (+) Absolute change in handedness* | 0.09 | **-132.24** | **0.28** |
| Model 2: (+) Absolute change in handedness*, (+) Index A of directional bradykinesia for unaffected hand | 0.13 | -119.08 | 0.29 |
| Model 3: (+) Oedema difference**, (-) Index B of directional hypokinesia for affected hand**, (-) Delta finger-to-palm distance ratio* | 0.25 | -105.79 | 0.30 |
| Model 4: (+) Oedema difference**, (-) Index B of directional hypokinesia for affected hand*, (-) Delta finger-to-palm distance ratio*, (+) Mood disturbance | 0.27 | -105.68 | 0.30 |
| Model 5: (+) Oedema difference**, (-) Index B of directional hypokinesia for affected hand**, (+) Mental Number Line Bisection score*, (+) Absolute change in handedness, (-) Delta finger-to-palm distance ratio | 0.32 | -107.60 | 0.35 |
| **Change in CRPS severity<sup>†</sup>** |  |  |  |
| Model 1: (+) Current pain intensity** | 0.13 | -50.21 | 0.58 |
| Model 2: (+) Current pain intensity**, (-) Index B of directional bradykinesia for unaffected hand* | 0.23 | -48.39 | 0.60 |
| Model 3: (+) Current pain intensity***, (+) Oedema difference*, (+) Hand laterality recognition accuracy index* | 0.25 | **-55.52** | **0.54** |
| Model 4: (+) Allodynia on affected limb*, (-) Index B of directional bradykinesia for unaffected hand*, (-) Index B of directional hypokinesia for unaffected limb*, (+) disease duration | 0.21 | -45.66 | 0.62 |
| Model 5: (-) Index B of directional bradykinesia for unaffected hand*, (-) Index B of directional hypokinesia for unaffected limb, (+) Allodynia on affected limb, (+) Disease duration, (+) Body perception disturbance score | 0.21 | -44.84 | 0.62 |

<sup>†</sup> Predicted outcomes were quantified as individual regression slopes based on pain intensity ratings throughout RS1-RS4 and LTFU1-LTFU2, and CRPS severity scores throughout RS1-RS4 (negative slopes indicate reductions in pain/CRPS severity).
\* $p < .05$ , \*\* $p < .01$ , \*\*\* $p < .001$ indicate significant predictors; (+), positive predictor; (-), negative predictor.
Adj. $R^2$ , adjusted R-squared; AIC, Akaike Information Criterion; CV, cross-validation error.
Figures in bold indicate the lowest AIC and CV.

## 4. Discussion

The results from this double-blind, randomized, sham-controlled trial do not support the effectiveness of PA treatment for upper-limb CRPS-I. First, we found no evidence that two weeks of twice-daily PA treatment performed with the affected arm reduced the primary outcomes of current pain intensity or symptom severity more than sham treatment in long-standing CRPS. Second, we found no evidence that PA affected the secondary outcomes of self-reported CRPS-related and psychological functioning; sensory, motor, and autonomic signs; or spatial cognition, motor function, and body representation.

Our findings contradict the conclusions of previous studies that PA could relieve pain and other CRPS symptoms. In the first of these, two weeks of once-daily PA training resulted in 50% pain relief, and reduced oedema and skin discoloration in five people with CRPS [100]. In the second study, three weeks of daily PA effectively resolved one patient’s pain, reduced autonomic symptoms, and improved motor function [9]. In the third study, four days of twice-daily PA resulted in 36% pain relief in seven people with CRPS [12]. In the two latter studies, its effects on pain were maintained for up to two weeks after discontinuing the treatment. While addressing the limitations of these preliminary small-sample, uncontrolled, unblinded studies, our robust trial showed no evidence of any benefits of PA for CRPS beyond those of a control treatment. A small reduction in pain intensity immediately following PA (13% reduction) was not significantly greater than after sham treatment (3%). Similarly, there was an overall reduction in CRPS severity immediately after treatment that persisted for four weeks, but was present in both PA (7%) and sham (8%) treatment groups. Although a lack of evidence for superiority of PA relative to sham treatment does not prove their equivalence, the effect sizes of any differences were negligible to small. Consistent across per-protocol and intention-to-treat analyses, there is no evidence that PA is any more effective than sham treatment for CRPS.

The decrease in CRPS severity across both treatment groups could be explained by a placebo effect and/or general benefits of moving the affected limb. Meta-analyses of clinical trials found that placebo response can correspond to an 1.84-point immediate post-treatment reduction in CRPS pain [60], or a 0.65-point reduction in chronic pain generally (on a 0-10 scale) [39]. This effect might also be responsible for the reduction in CRPS severity in our trial. Increased movement of the affected limb is a likely alternative explanation, because all participants performed the pointing task with their affected hand, regardless of the treatment condition. Physical exercise is a core pillar of CRPS management [28], and this additional daily activity might have been sufficient to reduce CRPS severity. It is unlikely that the observed changes were due to natural recovery, which might occur within the first year from diagnosis [1], as participants were on average diagnosed with CRPS for five years. Disease duration was also unrelated to changes in pain intensity or CRPS severity (Supplemental Figure S3). Regression to the mean cannot fully account for the decrease in CRPS severity, as no changes occurred over the baseline period. Overall, our findings reinforce the importance of including control treatment arms in pain rehabilitation studies, and the role of active movement in managing long-standing CRPS.

We address three potential reasons why we did not find the hypothesised effects of PA on clinical outcomes: (1) non-central pathophysiology of CRPS, (2) absence of neuropsychological symptoms, and (3) trial limitations. First, because PA targets neuropsychological deficits, it would possibly be most appropriate for a subset of individuals who predominately show signs of central neuroplasticity (compared to peripheral inflammation) [5,16]. However, post-hoc classification of participants into central or peripheral phenotypes and follow-up exploratory subgroup analysis did not reveal different responses to PA versus sham treatment (Supplemental Table S1 and Text S3).

Second, it is possible that we found no effect of PA on participants’ spatial cognition or body representation because, in contrast to previous findings [11,21,25,27,76,84,85,94,99], they did not have any systematic deficits on baseline experimental measures of spatial cognition and body representation. One hypothesised mechanism of the apparent benefits of PA in previous CRPS studies is that it reduces pain by correcting the “neglect-like” bias away from the affected side. A potential second mechanism is based on the proposal that distorted body representation gives rise to discrepancies between anticipated and actual consequences of movement, which cause or exacerbate pain in conditions such as CRPS [8,38,61,62]. The transient sensorimotor incongruence introduced by wearing prisms is thought to provide an error signal that triggers normalisation of body representation and sensorimotor integration [9,100]. On average, our participants showed balanced distributions of spatial attention and spatial representations, no systematic slowing of movements directed towards the affected side, and unimpaired laterality recognition of images of affected hands at baseline (see Table 2 and [32]). Cognitive after-effects of PA have been shown to depend on baseline spatial bias [14,29,44]. Therefore, if altering spatial cognition and/or body representation were integral mechanisms through which PA reduces CRPS symptoms, the absence of pre-existing neuropsychological deficits could preclude any effects of PA on the primary clinical outcomes. However, we dismiss this explanation, based on the following reasons. In the follow-up exploratory analyses, we found no relationships between the extent of baseline spatial or body representation deficits and changes in the primary outcomes over the treatment period (Supplemental Figures S4 and S5). We also found no evidence that PA benefitted subgroups of individuals who did present with “neglect-like” symptoms or distorted representation of the affected limb (Supplemental Text S3). Furthermore, Christophe et al. [12] reported reduced CRPS pain after PA in the absence of any baseline spatial deficits, and without any effect on spatial cognition or motor control. Finally, Sumitani et al. [100] found a significant reduction in pain post-treatment, and a simultaneous shift in the coding of external spatial information relative to the body *away from* the affected side (i.e. direction opposite to the expected PA spatial after-effects). Since these previous studies [12,100] had no control treatment arms, the apparent benefits of PA could be due to other non-specific factors. Nonetheless, overall it seems that response to PA treatment is unrelated to “neglect-like” spatial bias or body representation distortion.

Third, we considered several limitations of our study that might explain why we did not find the hypothesised effects of PA. Since we tested a protocol of PA that could realistically be integrated into CRPS management as a self-administered, home-based treatment, we cannot rule out compliance violations. We relied solely upon participants’ self-reported adherence, therefore the lack of apparent difference between the effects of PA and sham treatment could be due to deviations from the instructed treatment protocol. However, previous CRPS studies reported symptom improvement following less frequent [100], fewer [12], and home-based [9] PA sessions. PA protocols similar to ours using sufficiently strong prisms (10-20°) and 10 or more treatment sessions also found generalizable, long-lasting effects on hemispatial neglect [24,95]. Our use of home-based treatments meant that it was not feasible to confirm adaptation by measuring pointing after-affects. Yet previous studies using 50 pointing movements have shown that this is sufficient to create after-effects [9,90,100]. Overall, we do not consider that these limitations provide reason to doubt our findings that PA is not an effective treatment for long-standing CRPS. Nonetheless, greater confidence could be gained from a trial of supervised PA with a greater number of sessions, and confirmed adaptation. Similarly, acute patients, in whom symptoms are less established, might yet benefit from PA.

This longitudinal study allowed us to explore potential baseline predictors of CRPS progression over 10-30 weeks, regardless of treatment. Smaller change in hand preference since CRPS onset predicted greater reduction in pain intensity. Consistent with the learned non-use hypothesis [80], underutilization of CRPS-affected limb and compensatory use of the unaffected extremity might maintain CRPS symptoms and hinder recovery. Overall reduction in CRPS severity was predicted by smaller pain intensity and oedema of the affected limb, suggesting that people with milder symptoms are likely to improve more. Individuals who were better at recognising images of affected relative to unaffected hands also achieved greater reduction in CRPS severity. Body perception disturbance was previously linked to longer CRPS duration and more severe sensory and motor signs of CRPS [46,50,105]. Our findings that less distorted representation and maintained use of the affected limb predict greater symptom improvement support multidisciplinary pain management approaches, which aim to normalise body representation and foster active movement [28,73,74]. These interpretations are, however, tentative, as the analyses were exploratory and the abovementioned factors explained only 9% and 25% of variance in the overall changes in pain intensity and CRPS severity, respectively.

We conclude that there is no evidence that PA reduces pain and other symptoms more than sham treatment in long-standing CRPS. The benefits of PA reported in previous studies are likely due to the placebo effect, greater movement of the affected limb, regression to the mean, and/or natural recovery.

## Data Availability

Anonymised participant-level data generated during the current trial (https://osf.io/ba6fq/), digital study materials (training protocol and video, PsychoPy experiment files and stimuli; https://osf.io/7fk2v/), and analysis scripts (https://osf.io/w67rx/) are publicly available in an Open Science Framework repository.

## Acknowledgements

The study was supported by a grant from the Reflex Sympathetic Dystrophy Syndrome Association (RSDSA) awarded to JHB and MJP. The RSDSA approved the design of the study. The funders had no role in data collection and analysis, decision to publish, or preparation of the manuscript. We thank the individuals with CRPS for taking part in the trial, and the CRPS-UK Research Network, Dr Nicholas Shenker, Professor Candida McCabe, and other health professionals for their assistance with participant recruitment. We further thank our research assistants (Sophie Brown, Louise Quaife, Phoebe Carlisle, Matthew Haywood, Elizabeth Nikitopoulou, Sanya Manglani, Abdulaziz Aldaughaither, Danesh Paul, Rajavee Arora, Josephine Pedder, and Olivia Cousins) for their help with data entry.

## Supplemental Digital Content

### Supplemental Digital Content

A: Supplemental Text describing data preparation procedures (Text S1), results of per-protocol outcome analyses (Text S2), and exploratory correlational and subgroup analyses in intention-to-treat sample (Text S3). B: Supplemental Tables illustrating individual participant characteristics (Table S1) and results of per-protocol analyses, that is, baseline characteristics of PA and sham treatment groups (Table S2), group average scores on secondary outcome measures across all time points (Table S3), results of ANOVAs on secondary outcome measures (Table S4), and results of linear mixed models analyses for secondary outcome measures (Table S5). C: Supplemental Figures illustrating individual pain and CRPS severity reduction scores over the treatment period (Figure S1), average daily logbook ratings of pain intensity, symptom interference, and range of movement (Figure S2), scatterplots of changes on the primary outcomes vs. CRPS duration (Figure S3), scatterplots of changes on the primary outcomes vs. baseline performance on tests of spatial cognition (Figure S4), and scatterplots of changes on the primary outcomes vs. baseline scores on tests of body representation (Figure S5) in intention-to-treat sample; and primary outcomes in per-protocol sample (Figure S6).

